# Modeling the waves of Covid-19

**DOI:** 10.1101/2021.06.15.21258969

**Authors:** Ivan Cherednik

## Abstract

The challenges with modeling the spread of Covid-19 are its power-type growth during the middle stages with the exponents depending on time, and the saturations mainly due to the protective measures, though weakening and partial destruction of the virus due to mutations is a consideration too. The two-phase solution we propose for the total number of detected cases of Covid-19 describes the actual curves in many countries almost with the accuracy of physics laws. Bessel functions play the key role in our approach. The differential equations we obtain are of universal type; they describe momentum risk-management in behavioral psychology, transient processes in invasion ecology, etc. Due to a very small number of parameters, namely, the initial transmission rate and the intensity of the hard and soft measures, we obtain a convincing explanation of the surprising uniformity of the spread in many different areas. This theory can be used for forecasting the epidemic spread, evaluating the efficiency of the protective measures and the vaccinations. For instance, the early projection for the 3rd wave in the USA was very exact. The data until summer 2021 for India, South Africa and UK are discussed.

## Introduction

The evidence is strong that the exponential growth of the total number of detected infections of Covid-19, denoted by *u*(*t*) in this work, can be detected only during short periods. This is in any countries and especially when the middle stages are considered. The corresponding curves are in fact of power type: *u*(*t*) ∼*Ct*^*c*^ in terms of the time *t* from the beginning of the current wave for some *C, c*. The parameters *c, C* heavily depend on time; the exponent *c* approaches 1 near the turning point of the spread, and the magnitude *C* becomes small near the saturation. Since the epidemic is far from over, the saturation here is of technical nature. Generally, it is followed by a period of modest linear-type growth of the total number of infections.

Methodologically, we consider epidemics as “invasions”, and focus on “transiencies”, momentum managing the epidemic in this context, which approach results in a very exact modeling of Covid-19. This is actually similar to [Has]: “The question of interest was the time course of the epidemic, rather than the final state, which is always one where the disease dies out”. The “predator-pray” system for us is when the protective measures (including self-imposed ones) play the role of “predator”, and the “pray” is the number of infections.

This is different from the `SIR`-type models, applicable mostly to the initial periods of exponential spread and to final stages of epidemics. The `SIR` model was suggested in early 20th century. Since then, it was developed, but the exponential growth until the herd immunity is approached remains its essential feature. As we will show, the asymptotic periodicity of Bessel functions is absolutely relevant here. Generally, Bessel function and processes are important in mathematics and physics, but they were not employed for epidemics and in invasion ecology as far as we know.

Our approach seems promising in ecology. More specifically, Bessel functions can presumably describe various continuous 2-species models. Following [Has], the discretization, different time-scales, 3-species models are natural further steps. See also [HL, LPP]. Generally, `basic hypergeometric functions` and their variants are expected to occur.

The discretization will be discussed only a little. We also do not consider in this paper the concept of Momentum Risk Taking from [Ch2], somewhat similar to Kahneman’s “thinking-fast”, which can be considered as a behavioral counterpart of the “transiencies” in ecology.

The most ambitious here are the expectations that the same ODE model the processes of `momentum` decision making in our brain, but this is very preliminary. The number of neurons involved in the “momentum” analysis of some event is restricted here by the “predator”, the expected allocation of (very limited) resources of our brain for this particular task. The asymptotic periodicity of Bessel functions sets here some limits. Generally, it is surprising that Bessel functions, invented by Daniel Bernoulli long ago, are not one of the main tools in mathematical theory of epidemics, ecology, behavioral science, and beyond. Hopefully this will change.

The usage of the basic and current `reproduction numbers` *R*_0_, *R* is common for epidemics. The basic one, *R*_0_, is defined as the initial average number of people infected by one person who contracted the virus; see [CJLP, Co, CD, DHB, He, HL]. However, *R* can be used only qualitatively for Covid-19 and other epidemics of power growth: the formula *u*(*t*) ∼ *const R*^*t*^ for the total number of infections will stop working very quickly and cannot be of real help for the forecasting the spread of epidemics without very significant corrections.

Even *R* close to 1 would quickly begin to contradict the actual growth of infections of Covid-19. However, it is not unusual when they were reported as 2, 0.7, or so in the middle stages of the waves of Covid-19; they are provided constantly by Robert Kox Institute (for Germany) and other centers. This is questionable to us.

One of the possibilities to adjust `SIR` to the power growth of Covid-19 and other epidemics of non-exponential type (there were many such), is to assume that *R* ∼ 1 and that it is `non-dominant`, i.e. to invoke the theory of `resonances`. This provides a polynomial growth of the total number of infections, but such models are unstable mathematically. Our modeling is different: a combination of the “local herd immunity” with the role of active management.

One of the most efficient protective measures is (and always was) self-restriction of our contacts. Here the disease control centers are supposed to provide current information on the spread of the epidemic. This is of course combined with quite a spectrum of other measures.

It was already intensively discussed in the literature that the herd immunity can influence the spread of Covid-19 well before it reaches the levels of 70% or so. See e.g. [BBT]. The protective measure play a significant role in this reduction. However their relaxation can result in the recurrence of the waves of infection.

We actually make the next step: gclaim that `local herd immunity` begins almost from day one and that it results in the power-type growth of the total number of infections: *u*(*t*) ∼ *t*^*c*^. Here the exponent *c* depends very much on the time from the beginning of the wave of infections, which is addressed in our approach via Bessel functions.

Our theory was posted in the middle of April, when the saturation of the spread was observed only in several countries; they were mostly in phase one, under mode (*A*) in our terminology. We also provided a variant for `phase two`, under mode (*B*), when the hard measures are significantly reduced. The (*B*)-mode system of ODE appeared really necessary for correct modeling the spread. Qualitatively, `phase two` is the switch to some less aggressive management due to relatively low numbers of daily infections, including restricting and self-restricting our contacts.

For the initial growth *u*(*t*) ∼∼ *t*^*c*^ of the number of detected cases, the second phase is described well by *u*(*t*) ∼∼*t*^*c/*2^ cos(*d* log(*t*)) for some *d*. The passage from the Bessel-type functions *u*(*t*) for phase 1 to such ones can be clearly seen in many countries. Though the Bessel-type *u*(*t*) worked well almost until the saturation in quite a few, including our forecast for the 3rd wave in the USA.

The spread of Covid-19 in the USA was mathematically quite a challenge for us; the results of our prior efforts are systematically reported in [Ch1]. The first wave in the USA went through several stages, more than with any other countries we considered. Our understanding is that it was so mostly because the hard protective measures were constantly relaxed in the USA on the first signs of improvements, well before the actual saturation. This is in contrast to Europe and several countries in Asia. It was somewhat similar in UK, but it eventually reached phase 2 and the saturation of its first wave.

The costs and consequences of hard measures, especially lockdowns, are huge. However the saturation due to the protective measures is of unstable nature and the recurrence of the epidemic is quite likely if they are reduced or abandoned. Our theory generally provides the way to evaluate the efficiency of protective measures and employ them properly, but this is quite a challenge even if advanced mathematical means are used. See e.g. [FRAF].

### Prior approaches

There is increasing number of works where the power growth of the total number of infections is considered for modeling Covid-19. Let us mention at least [Ch1, MBS, MH, TKH].

Let us begin with [CD] (well before Covid-19). An ambiguity with the definition and practical calculation of *R*_0_, *R* is discussed there: “It is reassuring to know, however, that the sign of *R*_0_−1 is independent of the decomposition used and that the prediction of exponential growth or decay is therefore correctly made by any of the counting schemes.” This is our impression too: the sign of *R*−1 is what is mainly used practically, not the exact value of *R* (calculated by some simple formulas). We note that the spread is mostly assumed of exponential type in [CD]. Let us quote: “As far as we know, little can be said in general about the exceptional case that *R*_0_ is not strictly dominant”.

In [MBS], the authors comment on the power growth of the spread of Covid-19: “the nature is full of surprises”. In [TKH]: “this new contamination regime is hard to explain by traditional models”. In our one: “power law of epidemics must be the starting point of any analysis if we want our mathematical models to be up to date”. See also [Ray] and works mentioned there concerning a potential usage of `small-world interaction network`, where individuals are assumed to contact (mostly) local neighbors and have occasional long-range connections.

In a different direction, paper [BBT] and some other works suggest that the levels of `herd immunity` sufficient to impact the spread of Covid-19 can be significantly lower than the “classical” 70% or so: as low as 40% in some areas due to the population heterogeneity. From this viewpoint, we make the next step in this direction, which seems quite natural. Our starting assumption is that `local herd immunity` shapes the spread from the very beginning of epidemics and quickly reduces its exponential growth to the power one. This is related to the concept of `small-world`.

#### Spatial modeling

This approach is actually similar to the one via “small-world”. The graph of contacts, especially geographically related ones, is the key for spatial modeling. For instance, Fig. 8 in [BP] shows the initial spread of Covid-19 in Germany, which is naturally related to the geographic locations. The authors change there the `SEIRD model` (susceptible-exposed-infected-recovered-deceased) to `SEIQRD` by adding the quarantined compartment. Obviously quarantines and travel restrictions are important to model epidemics. See also Fig. 2 from [KBLK] (Germany too). The population heterogeneity is an important consideration here.

The challenge is to produce `meso-scale` forecasts, with specific information and for some concrete locations. See e.g. [JCO] (for Texas). The total number of detected cases and other general data for the whole country are insufficient for those in charge of practical managing the epidemic. Partial differential reaction-diffusion equations are used in [JCO]; see also [VLAB].

Classical methods “least-squares”, “Bayes”, “*k* nearest neighbors”, and various statistical tools can be used for “meso-forecasts”. Deep machine learning is a possibility here (which we will not touch upon). *AI*-systems generally do not help much with theoretical understanding the processes. There are too many parameters and the uniqueness of the optimal ones is not granted. However they “almost always” provide satisfactory projections. ODE, PDE, SDE are not strictly necessary for machine forecasting, but they are generally in terms of meaningful parameters, which can be of importance theoretically.

Surprisingly, the dimension of the graph of contacts is basically sufficient for forecasting the number of infections in sufficiently large areas: states, regions. This is our parameter *c*; the total number of infections is then ∼*t*^*c*^ at some early stages. This may hold for small areas, and even for the `viral load` in infected individuals (at the middle stages), but this is a `macro-phenomenon` in this paper. The uniformity of the curves of total numbers of detected infections in so many so different countries is still mysterious to us, though we think that we found proper mathematical tools to address this.

#### Power growth

The main problem with modeling Covid-19 appeared actually not the power growth itself, “power law of epidemics”, a starting point for us. This alone is insufficient for forecasting. Understanding the saturation is the key challenge for modeling epidemics. However, we must note here that the power growth of the number of infections is not commonly accepted, in spite of ample evidence during the Covid-19 pandemic.

Practically, the exponent *c* and the corresponding scaling coefficient *C* heavily depend on the time passed from the beginning of the corresponding wave of the infection. An exact mathematical model of this time-dependence is necessary here, which was proposed in [Ch1] using the Bessel functions.

For instance, the approach of [MBS] to the power growth was of experimental kind. The corresponding exponents depend very much on the considered periods. So the data in Figure 1 in [MBS] and in similar papers mainly show `qualitatively` that the growth is no greater than power. However the latter is quite convincing!

**Figure 1.**
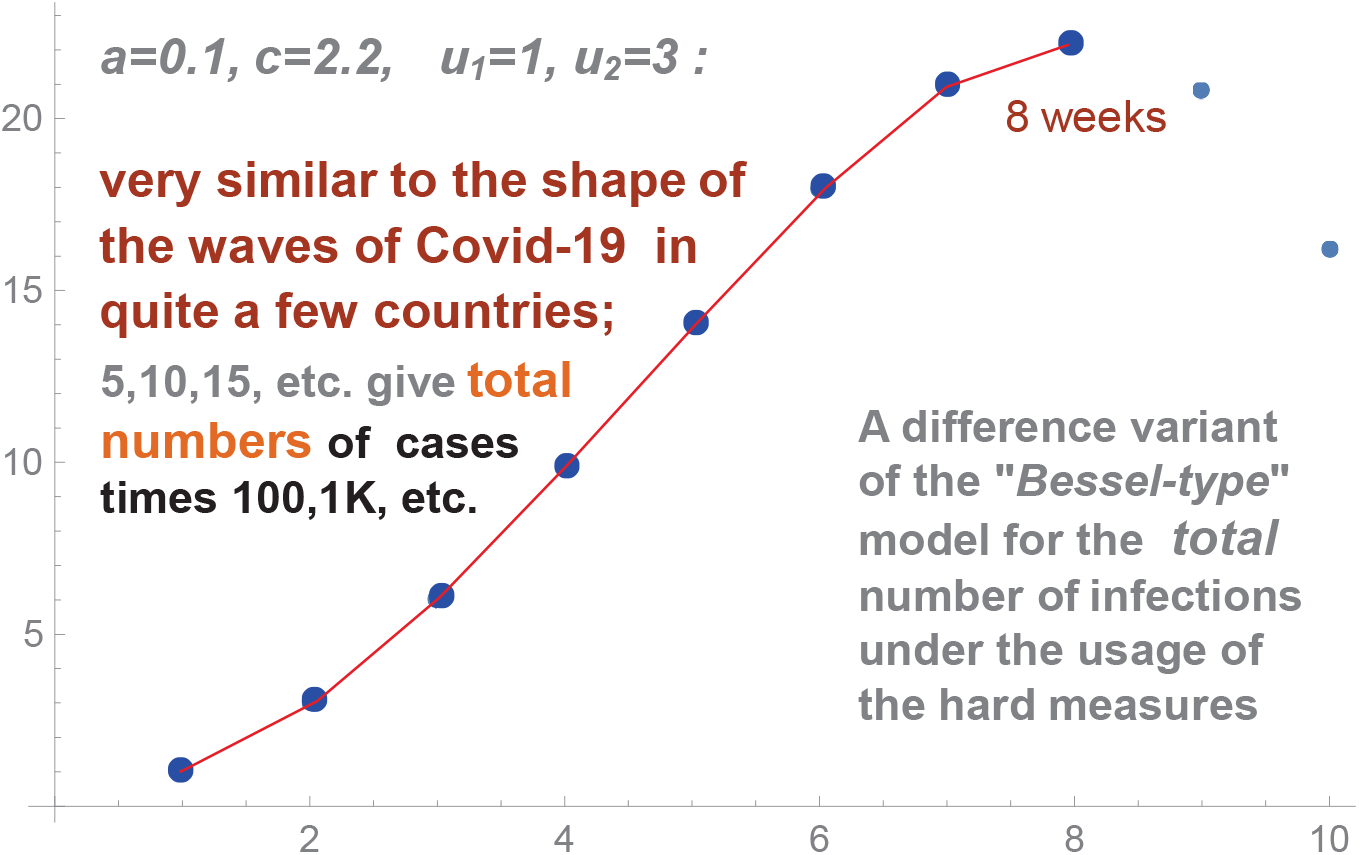
Discrete modeling the number of infections.

**Figure 2.**
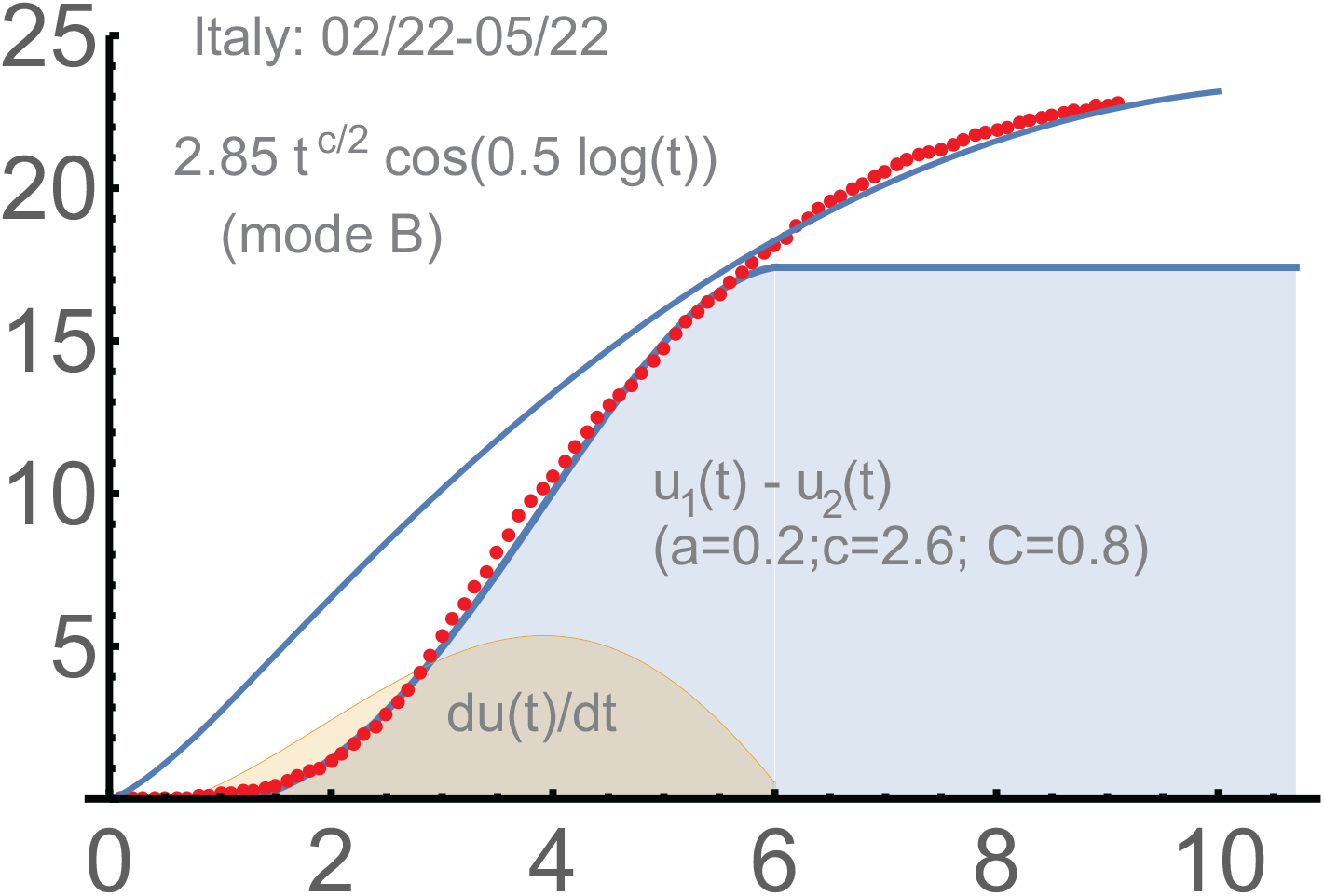
Italy: *c* = 2.6, *a* = 0.2, *d* = 0.5.

The parameter *c* we operate with is quite different from their exponents; it is for the whole period of the wave, even including the 2nd phase (where no Bessel functions are used). We mention that mathematically, the authors used the `SEIR` model (susceptible- exposed- infectious- removed), which does not result in the power growth; however, “small world” is mentioned there as a possibility.

Paper [TKH] is based on the Poissonian small-world network. This approach results in the linear growth (*c* ≃ 1) of the number of infections. The linear growth is indeed present indeed near the turning points of the curves of total numbers of infections, but it is far from linear anywhere apart from these middle portions. The explanation of the linear growth and the saturation in [TKH] is very different from what we proposed in [Ch1] and present in this paper.

Among many confirmations of the power growth of the total number of infections of Covid-19, the period 3/20-10/7 (2021) in India is very convincing; see Fig. 9. Here *u*(*t*) = *const t*^*c*^ for *c* = 3.65 is practically exact (almost without modifications) for the total number of detected cases in India for a very long period: for at least 5 months (!).

In this figure, the main parameters were determined on 08/03. This forecast was posted on 10/07 (2021); to expire on 11/06 (the maximum of the *u*-function). It matched very well the actual curve of detected cases. This was based on our usage of Bessel functions. Here and almost in any countries, some linear-type growth is expected after the top of the Bessel-type curve *u*(*t*), which can be seen in the graph for India after 11/06. In our theory, this period is described by the formula for mode (*B*); this is phases 2 in our terminology (for any waves).

### Saturation of Covid-19 waves

For us, the saturation, followed by some linear-type growth of the total number of infection, is due to protective measures, mostly the hard ones. They are those imposed by authorities in charge, but self-restrictions are equally important here. The key is `detection-isolation-tracing`, which includes closing the places where the spread of infection is the most likely. The societal cost of hard measures is huge, but they proved to reduced the spread efficiently. The vaccination is of this type too, a very hard measure by any standards.

Obviously, there are biological mechanisms that limit life of any virus strains. The mutations during consecutive transmissions mostly destroy or weaken it; at later stages of the waves of infection, those with weakened viruses (due to mutations) begin to dominate; the corresponding cases can be asymptomatic or mild, though the infected individuals create some immunity and transmit this weakened virus.

The mutations that make the virus stronger are very rare statistically, but they are present and can lead to new strains and new waves of the infection. With Covid-19, it must be like this too, but its proof-reading abilities reduce the total number of mutations and significantly increase the fraction of “constructive” ones and the creation of new strains. Importantly, it is relatively rare when individuals are infected by 2 different strains at the same time, though such cases can lead to dangerous transformations of the virus.

So, basically, if the strain weakened by the mutations begins to dominate then the current wave of infection (in this area or country) is on its way to the saturation. Much of course depends on the transmission strength of the strain. The more infectious ones spread the most.

These mechanisms and the role of the vaccination are present during later waves of Covid-19, near the end its current cycle. However it is not disputed that the saturation of the first waves of Covid-19 was not due to the them or herd immunity. The latter probably requires about 40%-60% (smaller than “classical” 70%) of all susceptible population to be infected and recovered [BBT]. It was far from these levels during the first waves. Thus, the saturation mechanisms of SIR-type models are not applicable here, at least for the 1st and 2nd waves.

The analysis of the second waves confirms the validity of our approach to modeling, based on the prime role of protective measures, mostly the hard ones. Recurrence of epidemics is quite frequent; see [HL]. However the second waves of Covid-19 began unusually quickly (for epidemics), sometimes even on the top of the unfinished first waves, as it happened in the USA. The relaxation of hard measures closer to the end of the first and further waves seems the only logical explanation of such sequences of waves.

In 2020, the summer vacations (and closed schools) in Western Europe were a clear instance of (broadly understood) hard measures. They did reduce the spread. However, at the end of August, the second waves began practically everywhere in Europe. Similarly, the number of new detected infections began to grow quickly (again) in the USA from the middle of September (2020).

The exponent “*c*”, which we call the `initial transmission rate`, appeared increasing from the first to the second waves in many countries. This parameter is one of the main in our theory; it reflects the virus transmissibility strength and initial the number of contacts in the area. Thus, by reducing the protective measures, *c* can be expected to be essentially back to its levels in the beginning of the epidemic, though with a tendency to increase. The increase of *c* from the 1st wave to the 2nd-3rd ones is related to the reduction of Covid-related restrictions (including our own behavior). The new strains of Covid-19 contributed too; its evolution is of obvious importance.

The second key parameter of our theory, the intensity *a* of protective measures, dropped `very significantly` for the second waves, which was expected. The intensity of the hard measures during the 1st wave was difficult to sustain during the next waves.

Qualitatively the duration of the wave is 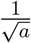; quantitatively, Bessel functions must be used here for exact modeling. So, mathematically, we essentially repeat the first waves, but now with significantly lower levels of hard measures, longer periods of intensive infections, and higher magnitudes of the curves of total numbers of detected infections. Though, the example of 2 waves in India is somewhat exceptional.

#### Later waves

The mechanisms restricted the duration of the waves become somewhat different at later stages, when the vaccination, herd immunity, and the self-destruction of the virus take effect. A striking similarity of the “delta-waves” in South Africa and UK (March-August, 2021), under the dominance of the Delta strain of Covid-19, indicates that there can be some `biological limits` for the durations of the waves, though of course much depends on the strain. These countries had very different situations (including the health care systems and vaccinations), but the corresponding curves of total number of infections are very similar. We will also note that in many countries the waves stopped when only some fraction of population was infected.

We think that, generally, `protective-restrictive mechanisms` are: `(a)`: protective measures by the authorities in charge, especially the hard ones, where “detection-isolation-tracing” of infected individuals is the key; `(b)`: self-imposed restrictions by the population, which are mostly based on the data provided by the authorities; `(c)`: herd immunity, the vaccination programs, and improvements of the treatment of the infected individuals, but they can be strain-specific; `(d)`: weakening the virus due to the mutations during chains of transmissions, though “proofreading” is of importance for Covid-19.

The main point of our ODE is applicable to all 4: the derivative of “preventions” is proportional to the current total number of infections. This means essentially that *N* preventions at the moment *t*_0_ (under (*a, b, c, d*), any of them) diminish the total number of infections at the moment *t* by (*t* − *t*_0_)*N* with some coefficient of proportionality. The preventions of any kind are proportional to the current number of cases.

### Power Law of Epidemics

With such complex processes as epidemics, there can be of course multiple factors contributing to the power growth, biological ones included [CLL]. The “justification” from [Ch1] goes as follows. First, we assume that infected people mostly transmit the disease to their (susceptible) neighbors, and that the population is distributed uniformly. The second assumption is that the wave of the infections expands linearly in a proper `graph of contacts`. The third assumption, the principle of `local herd immunity`, is that people “inside the infection zone” do not transmit the disease because they are surrounded by those already infected or recovered, i.e. the border of this zone mostly contributes to the spread of this disease. This readily gives that *u*(*t*) ∼ *t*^2^ or greater (in the absence of protective measures). Indeed the lowest *c* we observed was *c* = 2.2 (the 1st wave in the USA).

People from infected zones do shopping, travel, visit friends. So the higher dimensions are needed to imbed the graph of contacts into some ℝ^*N*^ providing that the `geometric distances` between points representing people are essentially the numbers of links between them, i.e. that these distances reflect the intensity of the contacts.

Upon this embedding, we assume the uniform distribution of the points in ℝ^*N*^ representing people, and the `linear spread` of the disease in ℝ^*N*^. This is basic physics. Then, indeed, *u*(*t*) ∼ *Ct*^*c*^, where *c* is the “dimension” of the image of this graph, a number from 2 to *N*.

Next, we represent such *u*(*t*) as a solution of the differential equation *du*(*t*)*/dt* = *cu*(*t*)*/t*. This is standard when we need to add “external forces”, which will be protective measures, more generally, mechanisms restricting the spread of the virus. We argued above that the exponential growth is generally unsustainable. However the power growth is unsustainable long term too. This will be “corrected” as follows.

### Adding protection

Combining the initial power growth of the total number of detected infections *u*(*t*) with the impact of protective measures we obtain the following two systems of differential equations:

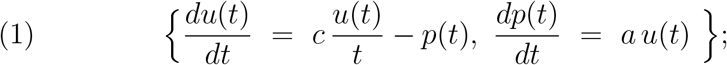

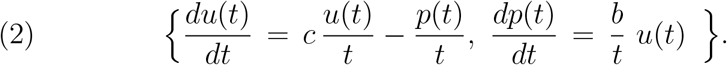

Here *t* is the time from the beginning of the intensive growth of infections, not always the very beginning of the corresponding wave of Covid-19 but sufficiently close to it. System (1) describes the impact of `hard measures` under the most aggressive response to the spread. The second system describes the impact of the `soft measures`: some travel restrictions, wearing the protective masks and social distancing are typical. We called these two modes (*A*) and (*B*) in [Ch1, Ch3].

When *a* = 0, *d* = 0, we obtain the power growth *u*(*t*) ∼ *Ct*^*c*^; so *c* can be measured experimentally during the initial stages of Covid-19 and is supposed to be the same for (1) and (2). Mostly it was in the range 2.2 ≤ *c* ≤ 2.8 (wave 1), but reached *c* = 4.5, 5.5 in Brazil and India.

There is a variant of these systems, when the second equation in is replaced by that from (2), called the transitional (*AB*)-mode in [Ch1]. It modeled reasonably the spread in the USA, UK, and Brazil, but the usage of (1) and (2) in our `two-phase solution` appeared sufficient for many countries without mode (*AB*).

The `protection function` *p*(*t*) for (1) is basically the number of prevented infections. More exactly, 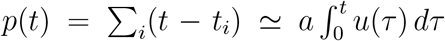, where the sum is over all infected individual isolated at the moments 0 *< t*_*i*_ *< t* for some constant *a*, the intensity of “isolations”. We assume that if not isolated, this group of people would contribute *p*(*t*) to *du*(*t*)*/dt* (the transmission rate is taken 1 for them). For (2), *p*(*t*) ∼ *(the number of infected people wearing the masks before t*), and, similarly, for other (self-)restrictions.

#### SIR-type modification

Let us touch upon the modification of system under the assumption that *u*(*t*) is bounded. Assuming that *u*(*t*) *<* 1, we multiply the right-hand side of the 1st equation by (1 − *u*(*t*)), which models the interaction of infected individual with the remaining (susceptible) ones. We obtain:

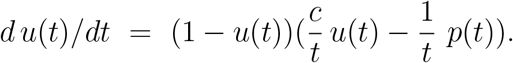

In the absence of *p*(*t*), it is a well-known *logistic equation*, with the following modification: the interaction is proportional here to 1*/t*.

The 2nd equation remains unchanged. The resulting system can be solved *numerically*. It is not clear whether the corresponding solutions are more relevant than those for the original system (2). For the following modification of the 2nd equation, this system can be readily integrated: *dp*(*t*)*/dt* = *b d u*(*t*)*/dt*. One has:

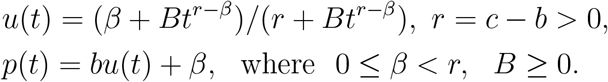

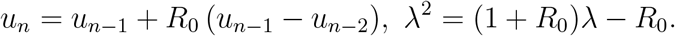

If *B >* 0, then *u*(0) = *β/r, p*(0) = *cβ/r, u*(∞) = 1, *p*(∞) = *b* + *β*.

### Related processes

Both systems are actually from [Ch2], where they were used to describe the dynamic of the (relative) stock prices *p*(*t*) under news driven `momentum trading`. The function *u*(*t*) there was the `news propagation` triggered by some event. It is of power growth in terms of time *t* passed from the event, but the exponent *c* is generally significantly smaller than 1, especially for short-term trading.

The arguments there were from `behavioral finance`. This is actually related; the behavioral aspects of epidemics are of obvious importance [St]. However financial news fades, and this happens quickly; this is very different for the spread of epidemics. System (1) described in [Ch2] `profit taking` in stock markets; the second one modeled the “usual” news-driven investing.

As a matter of fact, these two systems are of very general nature. For instance, they are supposed to occur in any `momentum risk taking`. This concept, MRT for short, is from [Ch2]; it is somewhat similar to Kahneman’s “thinking-fast” [Ka]. Managing epidemics on the basis of the current data is very much momentum. As in stock markets, people and authorities in charge must react promptly to any change of the situation. Another example there is `tree growth`, though there were no *p*(*t*) and the arguments were somewhat different.

It was expected in [Ch2, Ch1], though without biological evidence, that both systems of equations may describe real `neural processes` in our brain. Here *u*(*t*) is the number of neurons involved in the analysis of a particular event at the moment *t*, counted from the event, and *p*(*t*) is the expected importance of this event vs. other ones and the corresponding `expected` brain resources needed for its analysis. I.e. *p*(*t*) is basically the expected allocation of resources, which are very limited in our brain. We do not know much about the ways our brain work, but the confirmation of the power laws and related saturations are solid in the stock markets and, as we demonstrate, in epidemics.

We note that a significant part of [Ch2] is devoted to the `discretization`. Decision-making always requires some `action potentials`, i.e. it is discrete by its nature. With epidemics, the usage of ODE worked very well so far, though potentially the discretization can become important for our approach too.

### Toward discretization

We begin with some basics. *Let u*_*n*_ *be the total number of infections at the nth moment from the beginning of some epidemic*. Infected individuals transmit the disease mostly during some initial period, which we will make the unit of time. Let it be 1 week (indeed, about 1-2 weeks for Covid-19). The recurrence relation and the corresponding quadratic equation are:

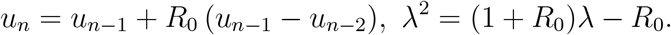

Here *R*_0_ is the initial `reproduction number`.

We obtain: 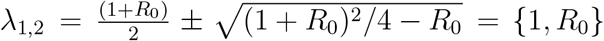. Thus, *u*_*n*_ = *C*_1_ + *C*_2_(*R*_0_)^*n*^ for some *C*_1,2_. If *C*_2_ ≠ 0 and *R*_0_ *>* 1, the growth of the total number of infections will be exponential.

This is unless the `herd immunity` is expected to be reached, when a difference version of `SIR` is needed. We will not address this here.

#### Power growth

Basically, there are 3 main mathematical possibilities to ensure a power growth:

i. the presence of “predators”, forces restricting the epidemic spread, for example, various protective measures,
ii. when *R* approaches 1, which results in “resonances” and can potentially provide some linear growth,
iii. when the “birth rate”, the transmission rate in this context, becomes inversely proportional to time.

Obviously (i) is applicable: we do fight epidemics. The resonances and linear growth occur when *R* ≈ 1, but this is unstable and does not seem of actual importance for modeling epidemics. We think, a combination of (i) and (iii) is the key in epidemics.

The spread of any disease is the growth of the “circles” of those infected; these are *combinatorial circles*, not geometric ones (in a map of the affected area). The contacts of infected people are not only with their immediate neighbors; people work, study, do shopping, travel. This is the concept of “small world” and the basis of spatial modeling. Our main assumption is that the rate of change of the radii of these circles can be expected constant. However they are not planar ones. The combinatorial distance is in the graph of connections (links) between people: “your co-worker” (the distance is 1), “a family member of your co-worker” (the distance is 2), and so on. The geographic connections are of course important here, but there are other links too.

The individuals at the frontier of such a circle transmit the disease the most because:

a. *they are the “latest” and therefore in their most infectious stages*,
b. *people inside the circle are “surrounded” by those with immunity*.

We assume that the infected people “inside the circle” contribute to the transmission of the disease significantly smaller than those at its boundary. Then the circle of infected people can be presented as a ball of dimension *c*; accordingly, the growth of *u*_*n*_ will be ∼*n*^*c*^. Here *c* can be any positive number, not only an integer. For instance, *c* ≈ 2 if our contacts are mostly with those who live near us, but *c* ≈ 3.65 in Fig. 9. We never saw countries with *c <* 2 during the 1st and the 2nd waves. Recall that *c*, the `initial transmission rate`, reflects the virus transmissibility strength and the number of contacts in the area at the beginning of the wave.

Disregarding “maturity”, a period when people are already infected but not infectious, the number of “newly infected people” is basically the area of the boundary of this “ball”, i.e. the area of the corresponding sphere. Presenting this number as *cu*_*n*−1_/(*n* − 1), we arrive at the following recurrence: 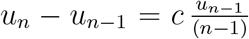. Asymptotically, *n*^*c*^ is a solution of this recurrence. Let us comment on this.

We have 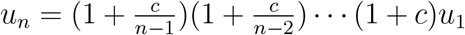. Therefore, 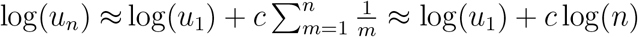 and 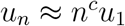. More exactly,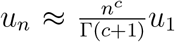 for the Gamma function Γ. When *c* = *r* for a positive integer 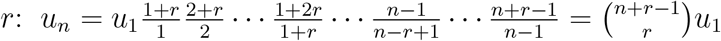.

#### Tree growth

We mention that the equation *u*_*n*_ = *u*_*n−*1_ +*u*_*n−*2_*/*(*n* − 2) describes reasonably middle stages of tree growth; here the timeunit is naturally 1 year and the “maturity” is set to 1. Its obvious solution is *u*_*n*_ = *n*. Under the initial conditions *u*_1_ = 0, *u*_2_ = 1, the corresponding solution tends to *n/e*. This solution is directly related to the `derangements` *D*_*n*_ in combinatorics: *u*_*n*_ = *D*_*n*_*/*(*n* 1)!.

The rationale for this model is that the volume of a tree is approximately proportional to *r*^3^, when the area of the root system is essentially proportional to *r*^2^, where *r* is the tree radius, which can be assumed to grow linearly. Thus the nutrition provided by the root system to one cubic unit of a tree is proportional to 1*/r* and to 1*/*(*time*). We omit the experimental support for this approach.

The following recurrence is convenient to model the saturation stage of tree growth: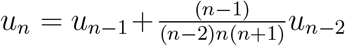. Its solution with the initial conditions *u*_1_ = 0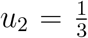, for 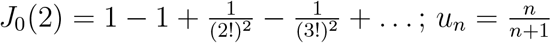 is another solution.

#### Adding protective measures

In the most aggressive variant of protective measures, we have:

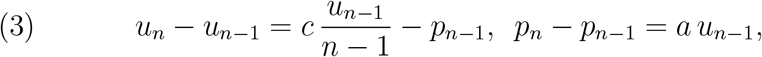

where *a* is the intensity of “hard measures”, *p*_*n*_ is essentially the number of people protected from the virus due to the measures until *n*. So we subtract the number of “preventions” from the total number of infections in the first equation.

This is parallel to the differential case; see (1). The main point here is that the “isolation” of one infected individual prevents the number of future virus transmissions roughly proportional to the time passed from this isolation.

The second equation is that the increase of *p*_*n*_, which is basically the increase of the number of preventions, is proportional to the current total number of infections, which is a very aggressive type of epidemic management.

Let us eliminate *p*_*n*_ from these relations. We obtain that 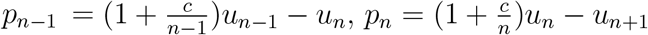, and

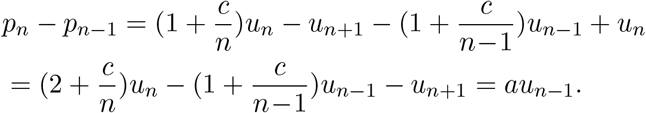

Finally, we obtain the recurrence relation:

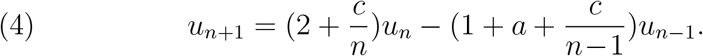

When *a* = 0, we have the solution *u*_*n*_ = *n* for any *c*. This is for *u*_1_ = 1, *u*_2_ = 2. For *a* = 0 and *arbitrary* initial conditions *u*_1_, *u*_2_, the “function” *u*_*n*_ is essentially proportional to *n*^*c*^, which follows from (3). When *c* = 0, this equation is not applicable: *p*_*n*_ = *u*_*n*_ − *u*_*n*+1_, the protective measures, cannot be negative. Recall that *u*_*n*_, the *total* number of cases, cannot decrease.

Fig. 1 is an example of the calculation with (4), where *a* = 0.1, *c* = 2.2, and we begin with *u*_1_ = 1, *u*_2_ = 3. We must stop at *n* = 8 (week 8), which is the saturation; the total number of cases cannot decrease. Near and after the maximum of *u*, a different recurrence must be used, which is a difference counterpart of (2), describing mode (*B*); see also *u*_*B*_(*t*) below. We will omit it.

### Two-phase solution

The solutions of (1) and (2) we need are

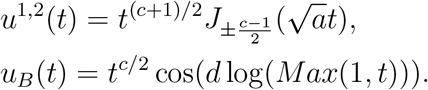

Here 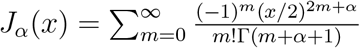 are `Bessel functions` of the first kind; [Wa] (Ch.3, S 3.1). The solution *u*^1^(*t*) is the main, though the second (non-dominant) solution *u*^2^(*t*) is important too. The function *u*_*B*_ is for 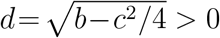; it will be used to model later stages of the waves of Covid-19.

Our `two-phase solution` is the usage of a proper linear combination of *u*^1,2^ for phase 1, till its saturation, and then the usage of *u*_*B*_ for phase 2. It proved to be quite exact for modeling the curves of total numbers of detected infections of Covid-19. For *t* ≈ 0: *u*^1^(*t*) ≈ *t*^*c*^ and *u*^2^(*t*) is approximately ∼*t*. I.e. *u*^1^ dominates; it is the key for forecasting.

The second fundamental solution of system 2 is with sin instead of cos. We note that when the protective measures are modest, we obtain *D* = *c*^2^*/*2 − *b >*0. The leading fundamental solution is *t*^*r*^ in this case with 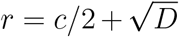, i.e. *t*^*c*^ in the beginning of the spread diminishes to *t*^*c/*2^ and then remains unchanged. This is of importance, but we will not touch the range *D >* 0 in this work.

The following examples of the 1st waves in 2020. mainly follow [Ch1].

#### Italy

`2/22-5/22`. Figure 2. The starting point was 2*/*22*/*2020, when the total number of infections was 17. We subtract this initial value here and below when calculating our dots, the total numbers of detected infections. One has:

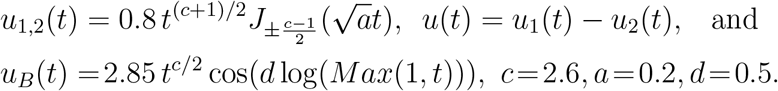

We use here both fundamental solutions *u*^1,2^(*t*) of system (1).

#### Germany

`3/07-5/22`. See Figure 3. We began with the initial number of total infections 684 (subtracted). This was approximately the moment when a systematic management began. One has:

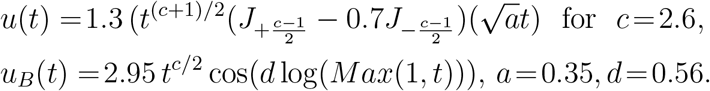

**Figure 3.**
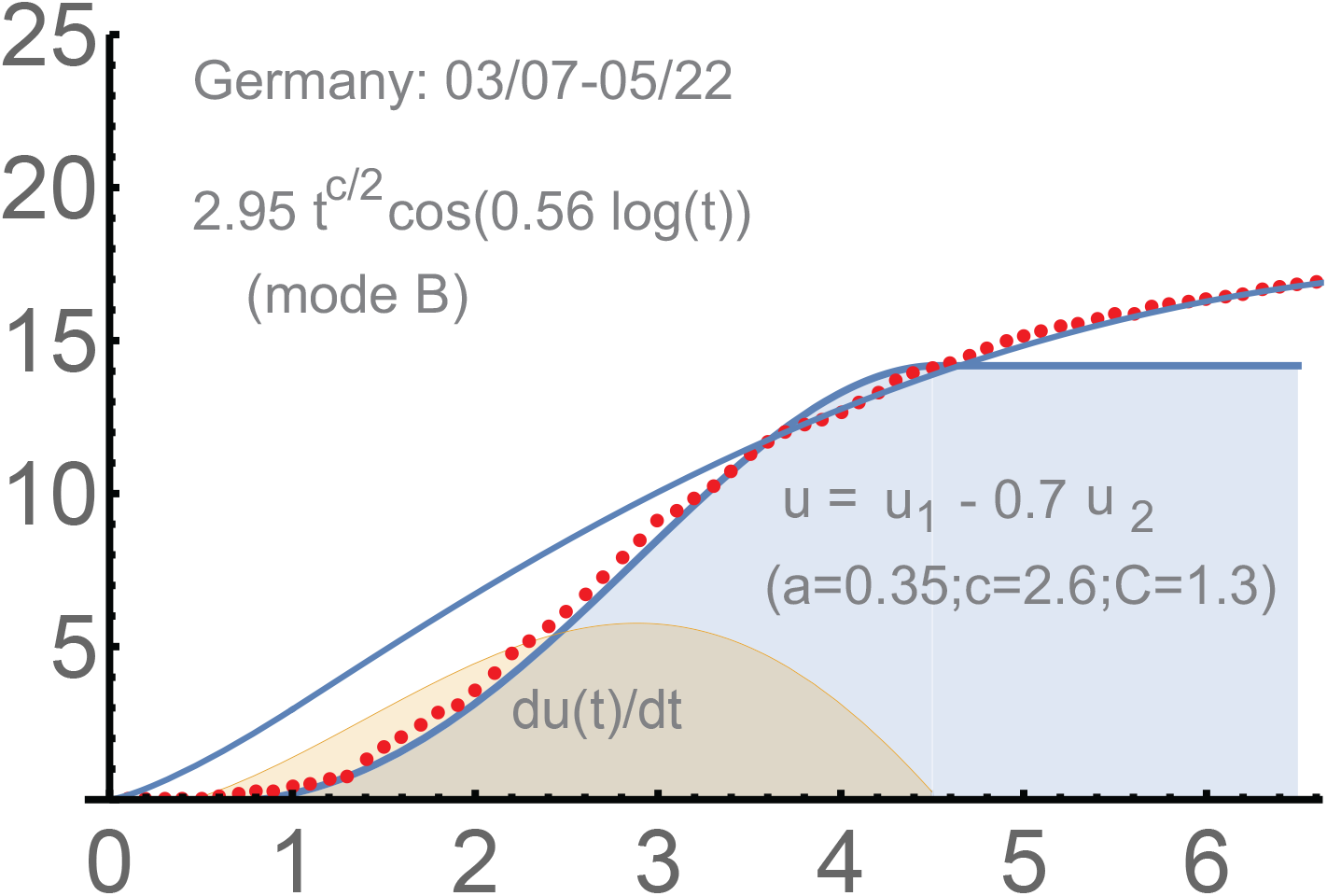
Germany: *c* = 2.6, *a* = 0.35, *d* = 0.56.

#### Japan

`3/20-5/22`. See Figure 4. There was some prior stage; we subtract 950, the total number of infections on March 20. The curve for Japan is not too smooth, which is not unusual. However it is managed well by our `2-phase solution` :

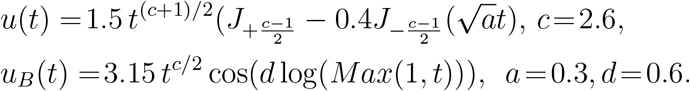

**Figure 4.**
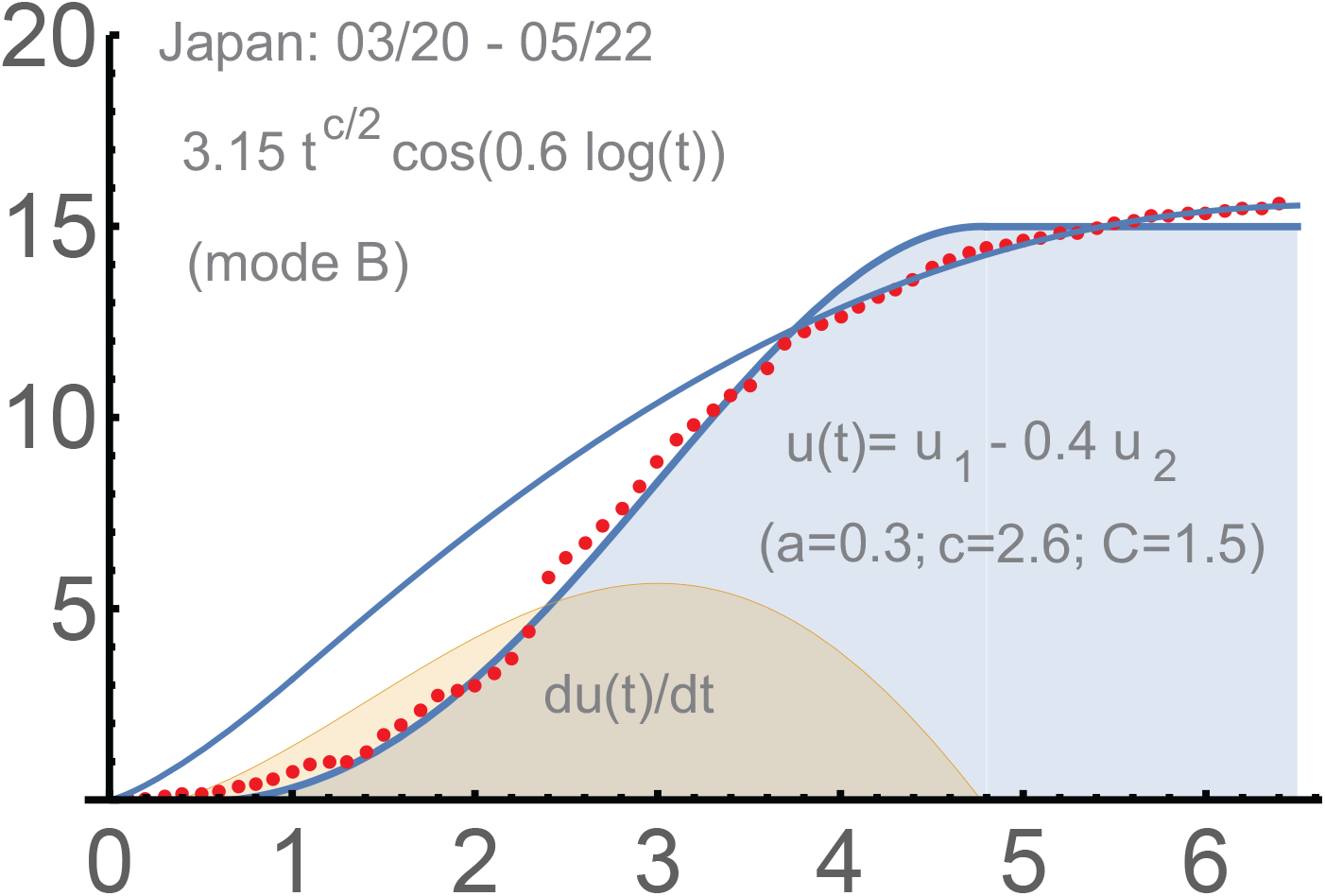
Japan: 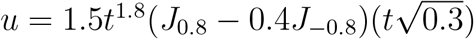

#### UK

`03/16-06/13`. This country was a challenge for us, though it “eventually” managed to reach phase 2. Actually the `red dots` for UK are modeled better with the transitional (*AB*)-mode. However, we prefer to stick to the “original” *u*(*t*) determined for the period till April 15. The two-phase solution is a combination of two phases separated by a linear period, about 10 days. See Figure 5. The formulas are:

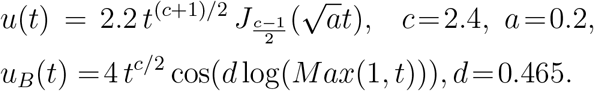

**Figure 5.**
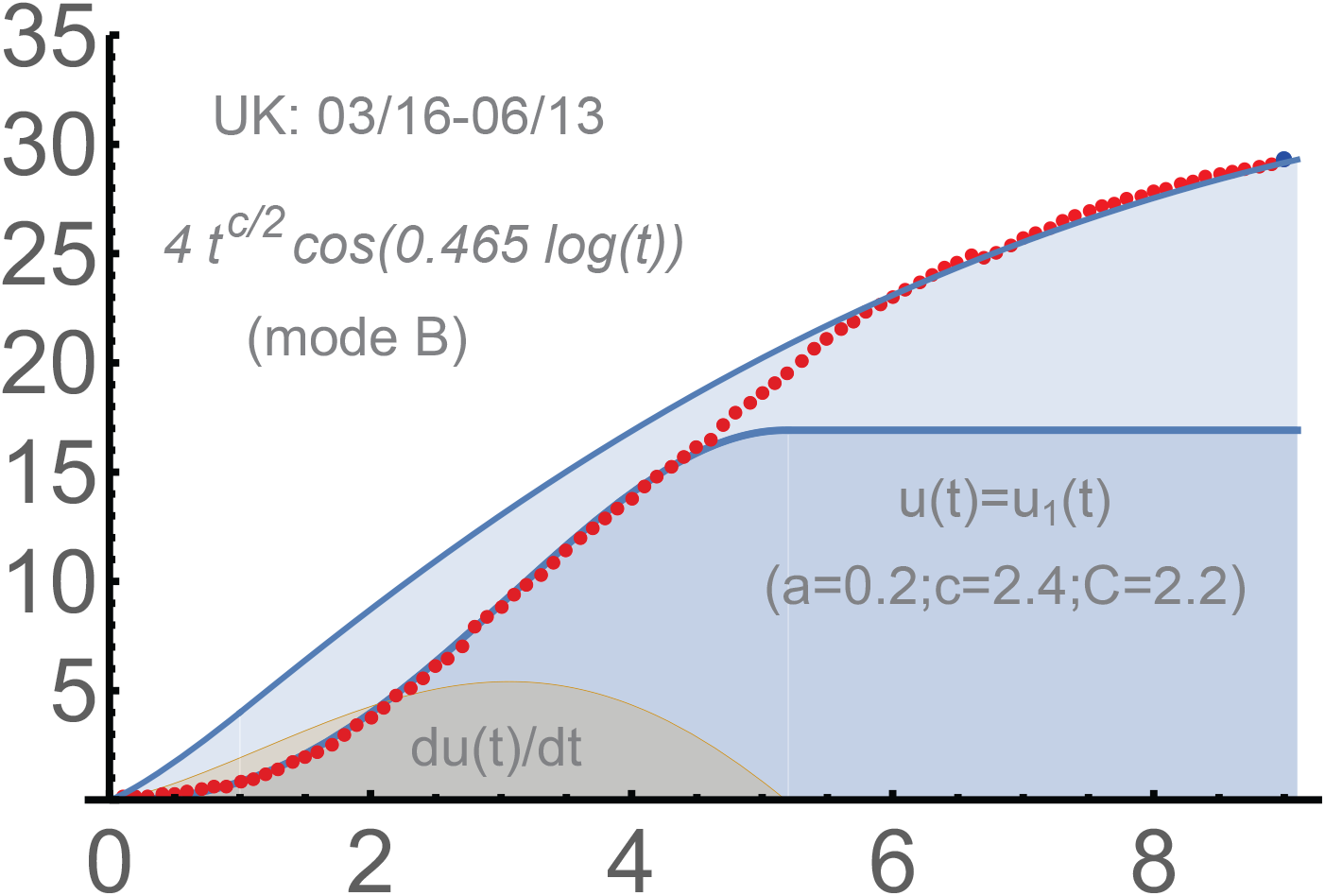
UK: 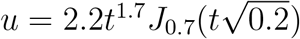

#### The Netherlands

`03/13-5/22`. The *u*-function here is with the same *a, c* as for UK. The parameter *d* = 0.54 is different from that for UK (*d* = 0.465). This could be expected; the process toward the saturation of phase 2 was slower for UK.

See Figure 6. The number of the total case was 383 on 3/13, the beginning of the intensive spread from our perspective. The usage of the dominant *u*^1^ appeared sufficient:

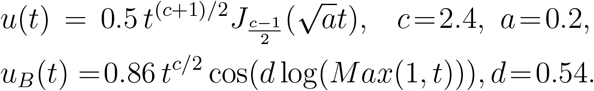

**Figure 6.**
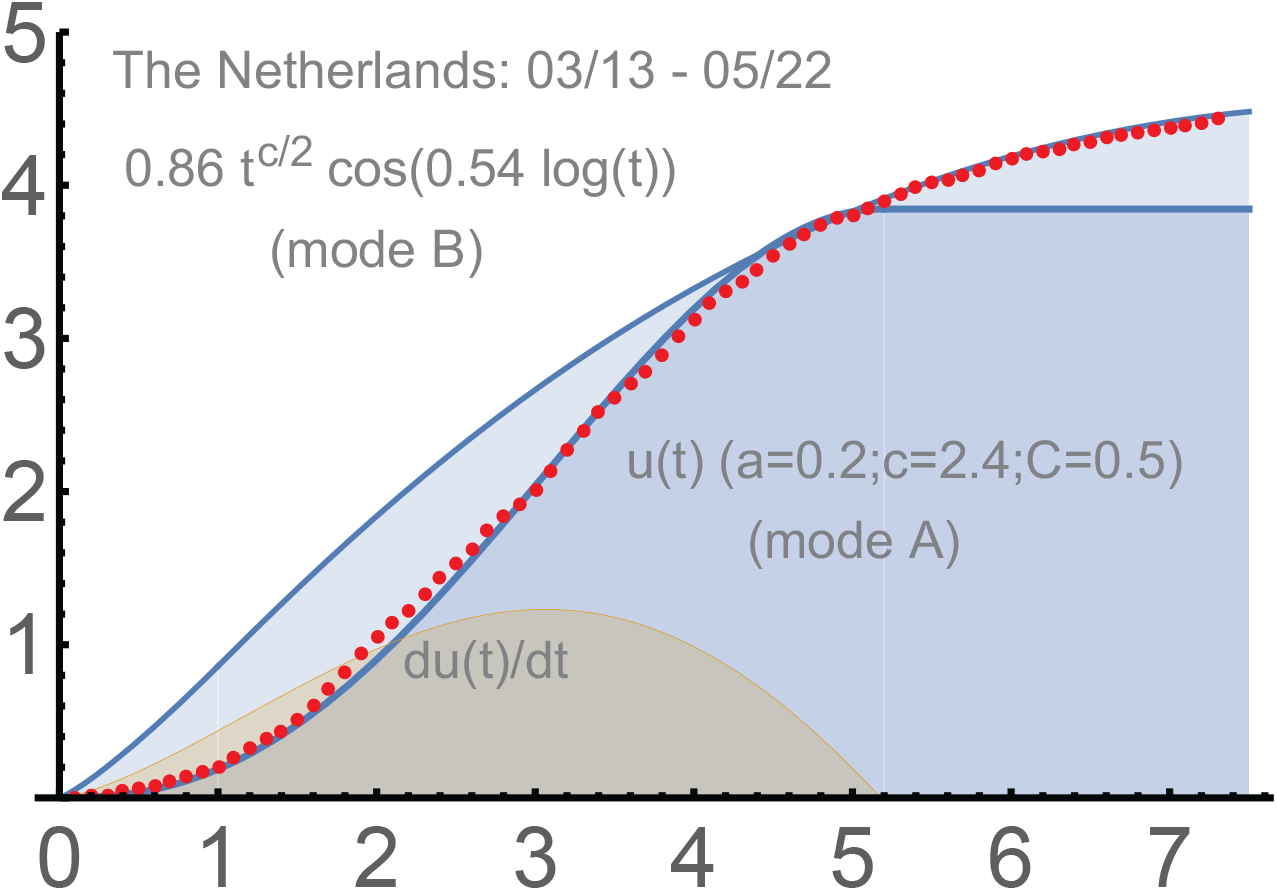
The Netherlands: 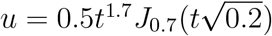.

### Further waves

The mathematical similarity of the second and 3rd waves to the first waves is very remarkable, a strong confirmation of our approach. The parameters *a, c, b* though change, which is generally in a quite understandable way. We will begin with the 2nd wave in the Netherlands, from 08/24/2020.

#### The Netherlands

`the 2nd wave, 2020`. The second waves were quite uniform in Western Europe. The Netherlands is convenient to demonstrate the evolution of our parameters, because the corresponding *u*-function does no involve too much of the second, non-dominating, Bessel-type solution. Generally, both are present.

In December of 2020, almost all Western Europe switched to a linear-type growth of the total number of detected infections. Later, the 3rd waves began there on top of the unfinished 2nd waves. The usage of protective measures became less stable, which can be partially due to the holiday season and the new strain, Alpha, of Covid-19, which was on its way to dominance. Within our modeling, the intensity parameters *a* diminished anywhere in Europe and the USA vs. those for the 1st waves, which is a clear indication for us of the significant reduction of the protective measures.

The similarity of Fig. 7 and Fig 6 is obvious. This is similar to the qualitative similarity of the 1st, the 2nd, and the 3rd waves in the USA to be discussed below.

**Figure 7.**
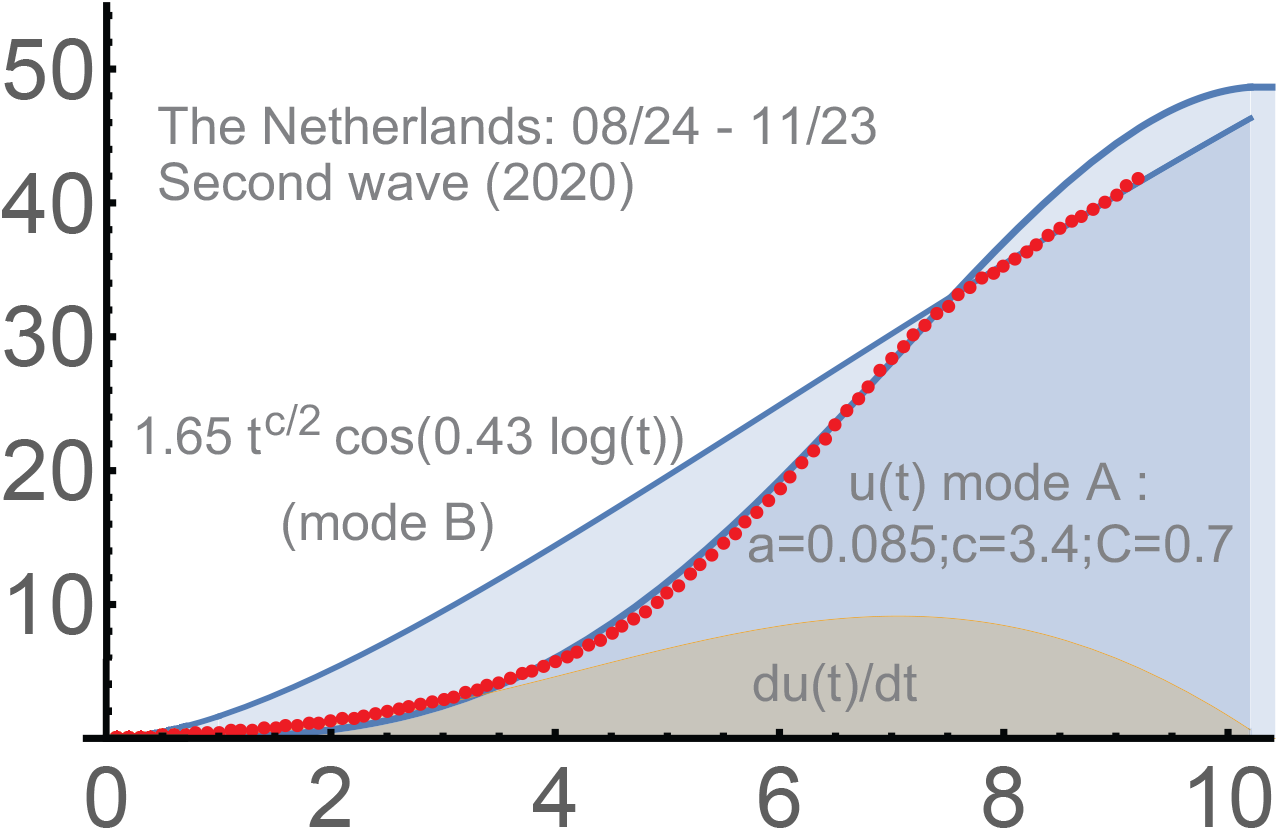
The 2nd wave in the Netherlands.

The parameter *c* significantly increased in the Netherlands: from 2.4 (the 1st wave) to 3.4 (the 2nd). The intensity of the hard measures understandably dropped: from 0.2 to 0.085. Such changes are actually common for the second waves in Europe. The parameter *d* = 0.43 diminished from 0.54. Our projection worked well until the beginning of December, actually until the pause between the 2nd and 3rd waves.

For the second wave in the Netherlands, one has:

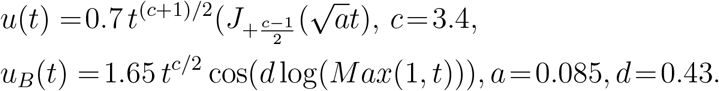

In Europe and the USA, there were significant fluctuations of the spread during the winter. Some hard measures were re-introduced in October-January. The vaccinations began in Spring, certainly very hard measures. This made and will make us closer to the herd immunity (for these particular strains). The self-imposed restrictions are of course of obvious importance here; their reduction can be one of the reasons for the increase of the initial transmission rates *c* during the 2nd waves vs. those for the 1st ones.

#### Japan

`the 4th wave (in process)`. It began in March, 2021. We take the period 03/15-5/25, 2021. The formula for *u*(*t*) is:

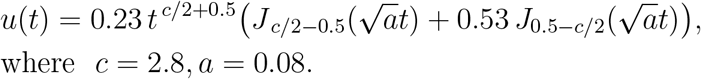

where *c* = 2.8, *a* = 0.08.

The tendency for *c* to somewhat increase and for *a* to drop is similar for that in other countries we considered. Here we did not create the control period; all dots are red. It is clear from our modeling that the 1st phase of the 4th wave approaches its end in Japan, and there is a switch in process to the 2nd phase.

#### The 1st wave in India

`3/20-10/07-11/20, 2020`. This country provide important mathematical patterns of the dynamic of the spread of Covid-19. The (clear) first wave was later than in quite a few countries, but it was with the greatest *c* we observed. The starting number of detected cases was 191, which was subtracted.

The power function 0.0125(*t* + 0.07)^3.65^ gave a surprisingly good approximation for more than 5 months; see Fig. 9. The parameter *c* began to decrease much faster in other countries. Obviously, the size of population is a factor here. This unusual stability the exponent can be also linked to a relatively low level of the active management in this country during the first wave. Of course the density of the population (which is huge in many parts of India) and the general number of contacts are very significant factors too. This is in India and anywhere.

**Figure 8.**
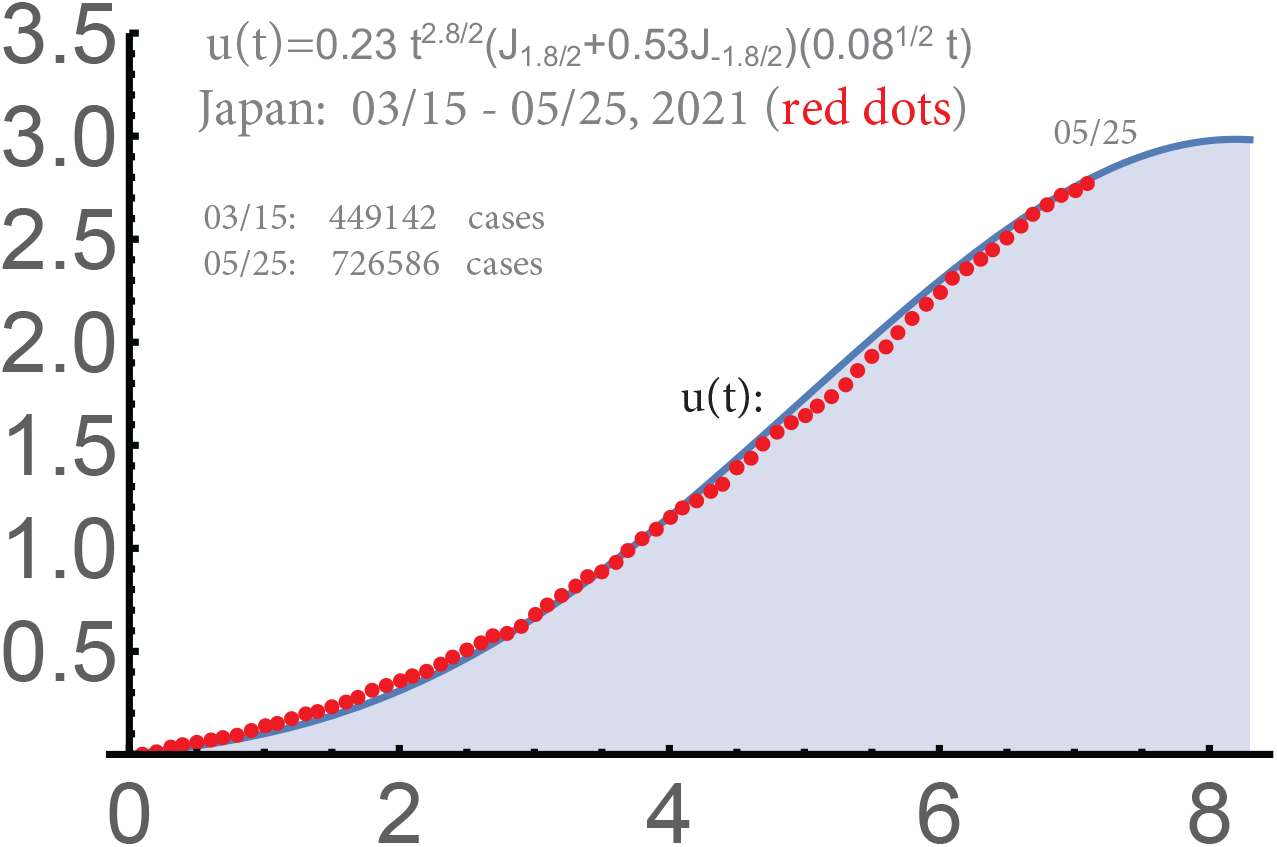
The 3rd wave in Japan.

**Figure 9.**
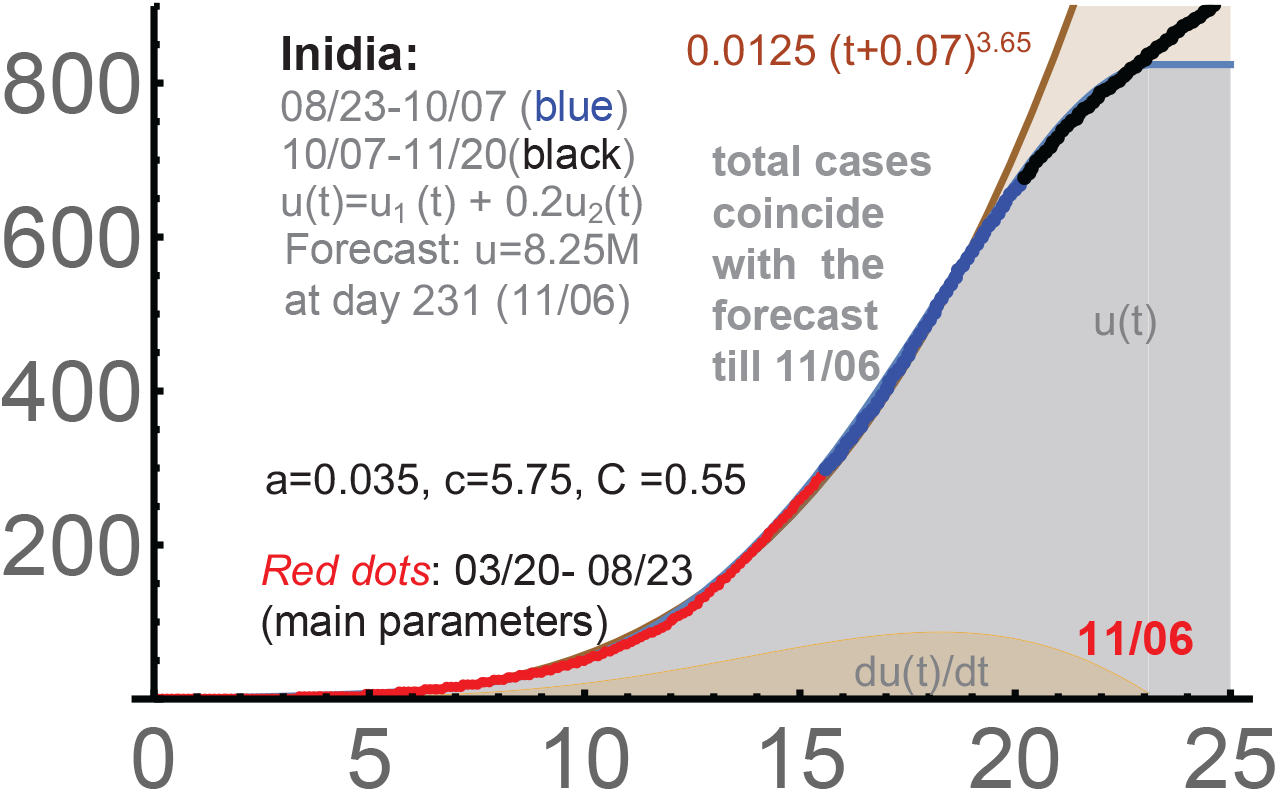
India: 3/20-10/7, *c* = 5.75, *a* = 0.035, *C* = 0.55.

Such a clear power growth is a very convincing argument in favor of the `power law of epidemics`. It is like this can be seen in all countries. It was mistakenly called exponential in some publications, which was of course not the case.

In Fig. 9, the parameters were determined around 08/03. Generally, the parameters determined before the `turning point` of the curves of total cases must be considered conditional, though we did it successfully in several cases before these points. The forecast posted on 10/07 was that the curve of the total number of detected infections would reach its technical saturation on November 6 with the number 8.25M of the cases. It matched the actual number of cases well.

As always, a linear-type growth (mode (*B*)) is expected after the top of the Bessel-type curve *u*(*t*), which can be seen in the graph. This period is described by *u*_*B*_. The parameter *c* is the same for phase 2 as it was for phase 1 (in the Bessel-type *u*(*t*)). Also, *u*_*B*_ begins from the starting point of the wave: “as if it were no 1st phase”.

We omit *u*_*B*_ for India. Let us mention here that our computer forecasting programs are for the 2nd phase only. They find the best *u*_*B*_ (from point 0) approximating the last 20 points. See below.

Here *y* =cases*/*10*K*; similarly, *y* is the total number of cases divided by proper powers of 10 in the other charts we will consider. Say, divided by 100*K* for the USA. The *x*-axis is always time in days from the beginning of the curve. The `red-blue-black dots` give the corresponding actual total numbers of the detected cases. The *u*-function for India is:

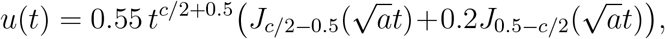

where *c* = 5.75, *a* = 0.035. It matched very well the actual numbers of cases till the middle of December (till the 2nd phase of the 1st wave).

#### The 2nd wave in India

`03/25-05/25, 2021`. The corresponding *u*-function is

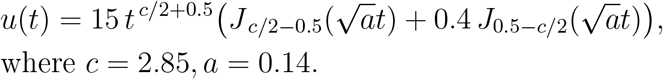

where *c* = 2.85, *a* = 0.14.

We note that the parameter *c*, which is the key, *became smaller* than during the 1st wave in India, though there is a somewhat greater contribution of the second solution *u*^2^, the term 0.4*J*_0.5*−c/*2_(). Generally, this term affects *C*, but does not influence much *c, a*. Its “role” is to adjust better the early stage of the wave. At later stages *J*_0.5*−c/*2_ becomes negative. This is why *C* must become greater in its presence with positive coefficient; this term was 0.2*J*_0.5*−c/*2_() for the 1st wave.

The coefficient *a* for the 2nd wave is quite similar to those for the 1st waves in Europe, better than those for further waves in Europe and the USA.Recall that the duration of the wave is qualitatively proportional to 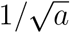; he duration of the 2nd wave in India was smaller than that of the 2nd-3rd waves in the USA and Europe.

The coefficient *c coincides* with that for the 3rd wave in the USA; it is much smaller than the one for the 1st wave in India, which was 5.75. Generally, this means that the population of India reacted faster (reduced the number of contacts) when the 2nd wave arrived. The self-imposed restrictions are very efficient *hard measures*. Biologically, the strain became more virulent. For instance, the likelihood increased dramatically for the whole family to become infected if one of its members is infectious vs. the 1st wave according to Indian medical officials.

We conclude that *much* greater number of detected cases is obviously related to the jump of the magnitude *C*. This is generally simply a scaling coefficient. However, when comparing different waves in *the same* country, it contains valuable information. Our theory results in the following output: between the waves, the virus in India acquired significantly greater ability to penetrate practically all strata and age groups of this very large and diverse country.

The control period was until 06/26/2021. The accuracy of the Bessel part of our 2-phase solution was very high. As it was with many countries, the first phase smoothly switched to the 2nd one (mode (*B*)). The formula for the 2nd phase is : *u*_*B*_(*t*) = 20.1*t*^*c/*2^ cos(0.53 log(*t*)).

In the USA and Europe, the changes between the first 2 waves were mostly due to the relaxation of the hard measure (including self-restrictions), i.e due to a very significant drop of the coefficient *a*. Some increase of *c* was of the same origin. However in India, *c* diminished and *a* increases, so the virus itself evolved: became a “broader one”. Generally, the changes of *c, a, C* can be used to analyze the trends in the virus’s evolution. Though they are very much linked to our response to the threat in our modeling.

### USA: waves 2 and 3

We use them heavily to adjust the forecasting component of our theory, especially the 3rd wave. The latter was used for (relatively) long-term forecasting, which appeared quite possible.

#### The 2nd wave in the USA

`06/16 - 9/12, 2020`. The two-phase solution worked well for the second wave in the USA. The accuracy is comparable with what we had above for the first waves in Japan, Italy, Germany, the Netherlands and UK. Upon subtracting 2.1*M*, the second phase matched well the following functions:

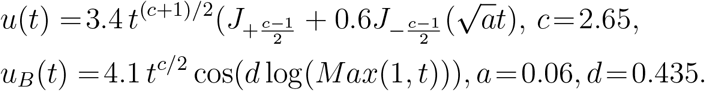

We note that the initial transmission rate was *c* = 2.2 for the USA during the first wave. The parameters *c, C* and 0.6 in the first formula were determined for the period marked by red dots; the black dots form a control period. See Figure 12. The projected saturation for *u*_*B*_ is given by the formula 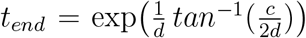. Numerically, *t*_*end*_ = 17.8463, which is 178 days from 06/16: December 11, 2020. Though this did not materialize since the USA entered the 3rd wave in the middle of September.

**Figure 10.**
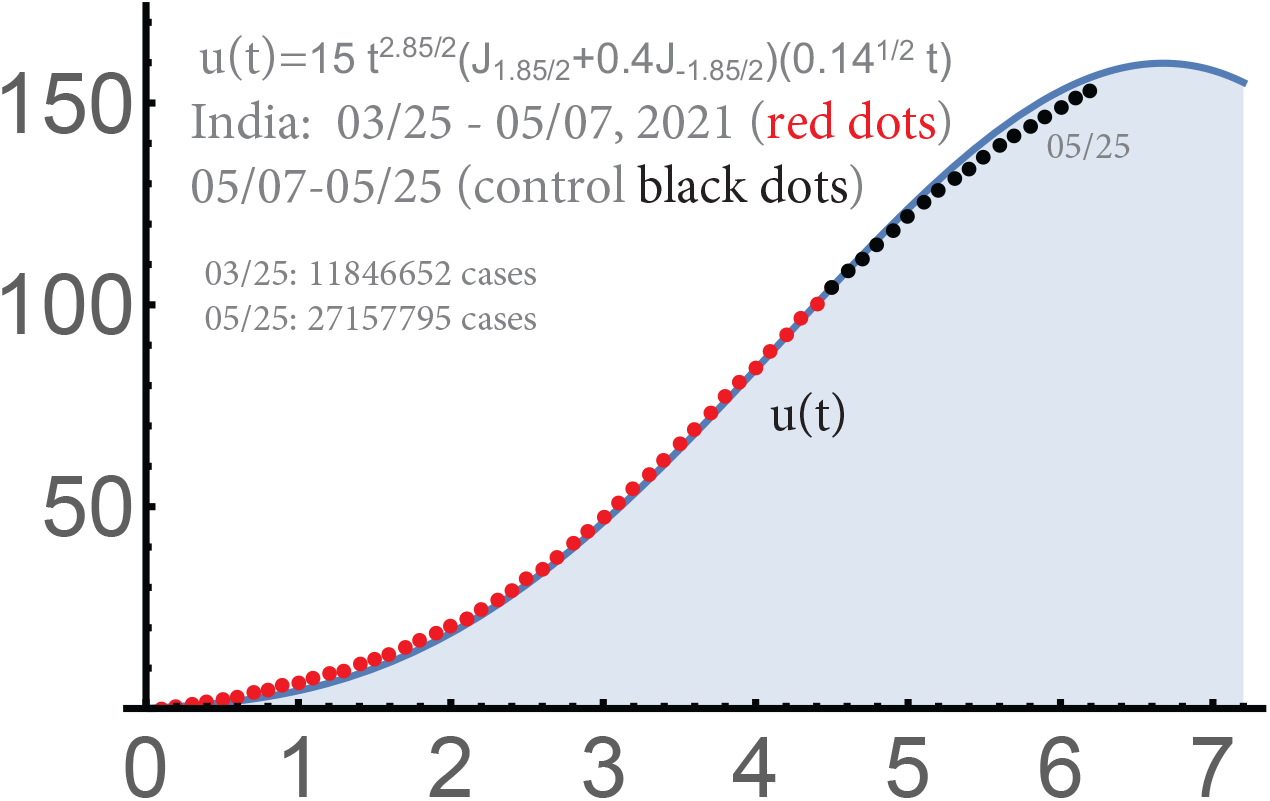
India: 2nd wave c = 2.85, *a = 0*.*14, C = 15*

**Figure 11.**
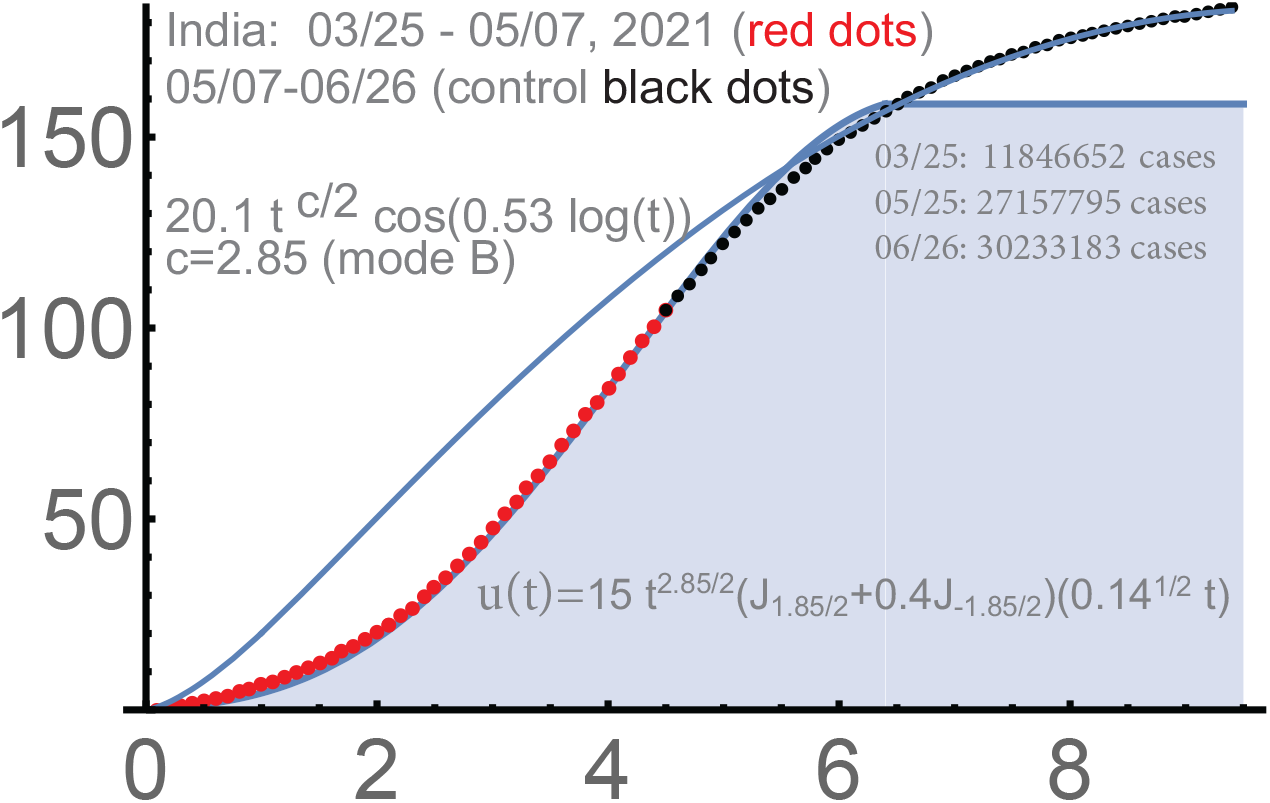
India: 2-phase solution until 06/26/2021

**Figure 12.**
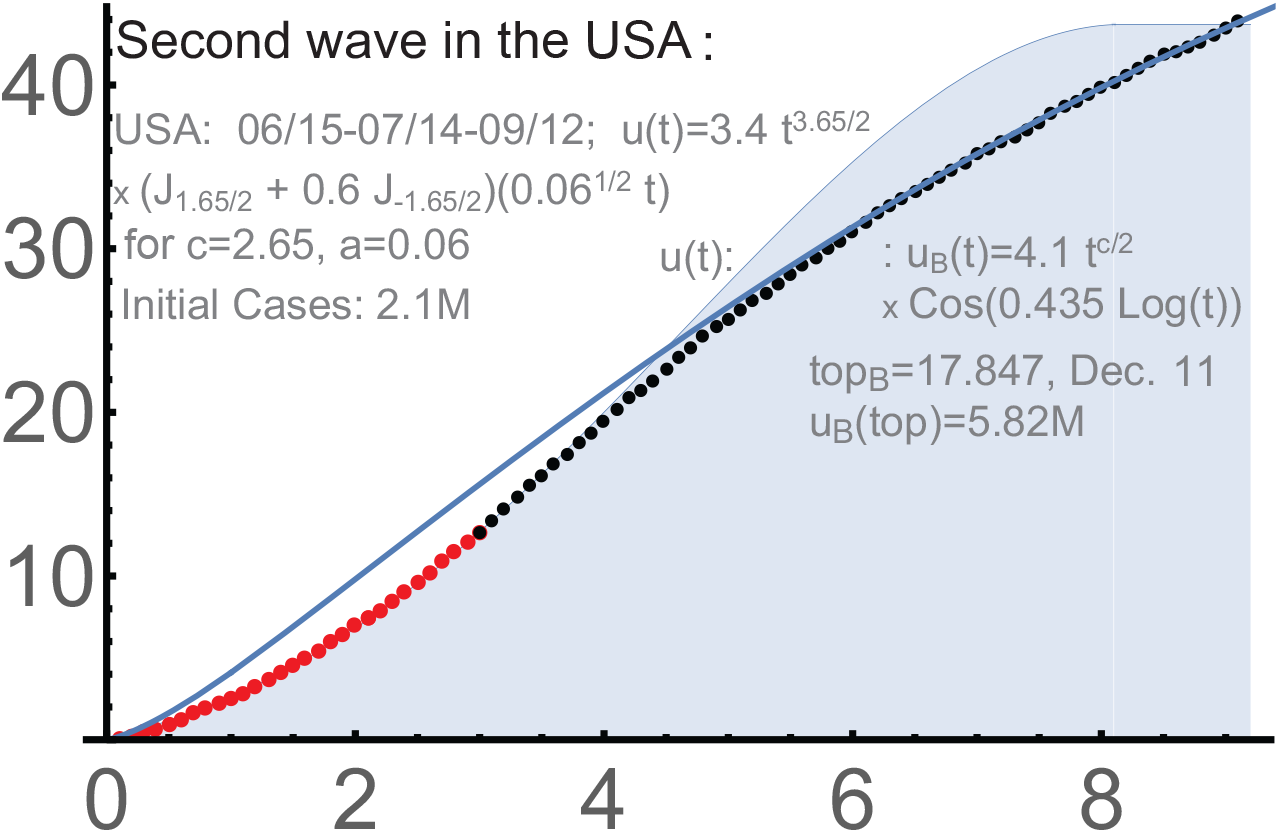
2-phase solution for the 2nd wave in the USA.

#### USA

`the 3rd wave, 2020-21`. This wave deserves a detailed analysis. It was used for long-term forecasting, which we will describe in consecutive steps, following the corresponding projections.

This wave was on top of the unfinished 2nd wave, so we subtract the starting total number of infections, which was about 6.9M on 9/24, when our curve begins. The red dots used to determine the parameters of *u*(*t*) were taken from 9/24 to 11/17/2020, when the 3rd wave in the USA still did not reach the turning point.

The 1st control period (black dots) was till 12/13. The match was already very good, See Fig. 13. However the accuracy became even better in March. Possibly some early signs of herd immunity contributed.

**Figure 13.**
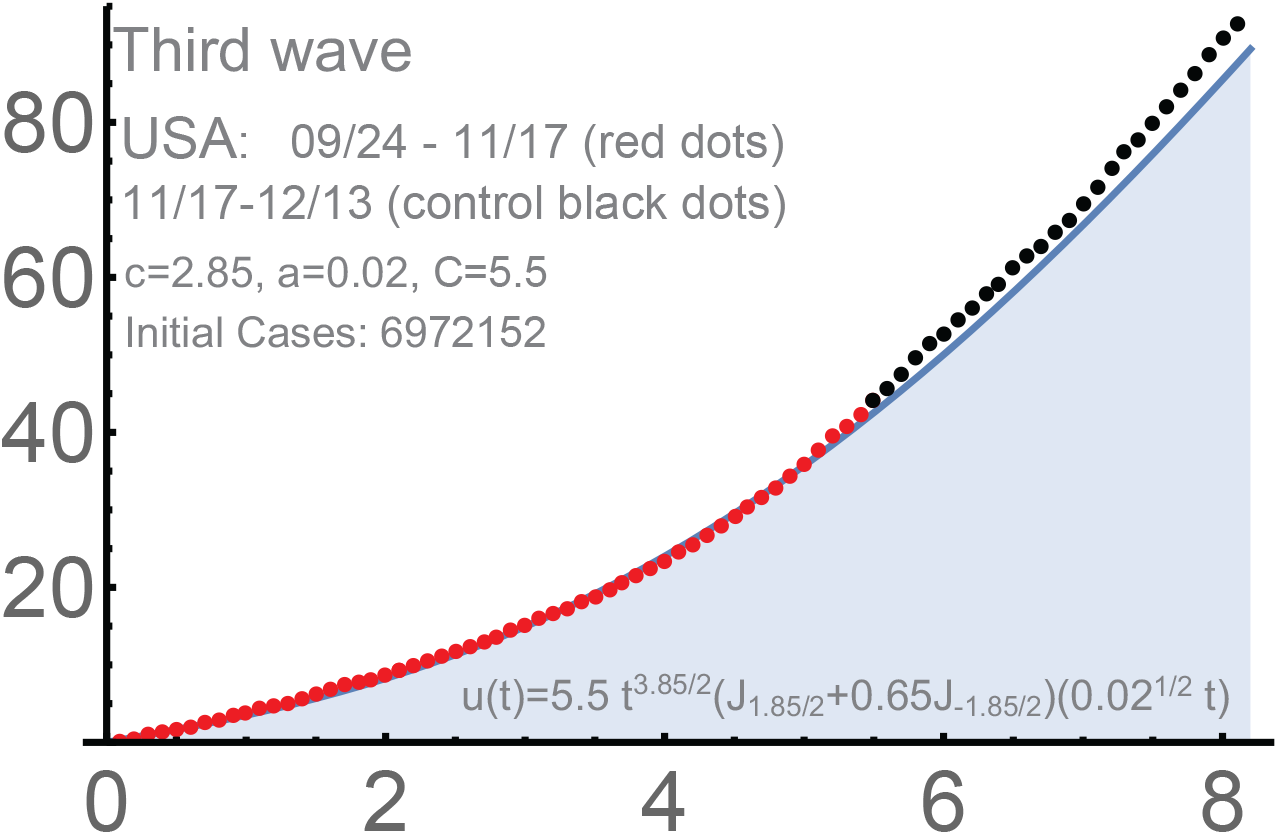
The 3rd wave in the USA.

The formula for *u*(*t*) for the 3rd wave was as follows:

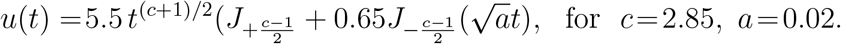

The new *c* increased from *c* = 2.65 for the 2nd wave to 2.85 in a way similar to the passage from the 1st wave to the 2nd. The parameter *a* significantly dropped from *a* = 0.06 for the 2nd wave to 0.02, 3-fold. We note that *a* = 0.06 was about 1*/*3*rd* of *a* = 0.2 for the 1st wave, so the same tendency persisted. Recall that the duration of phase (*A*) is qualitatively 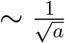.

The projection was 03/05/2021 for the saturation of the 3rd wave in the USA. It was supposed to be followed by some period of modest (essentially linear) growth under the (*B*)-mode (the second phase). This projection was posted on 11/17/2020; see Fig. 14. The match during the first control period (black dots) was good.

**Figure 14.**
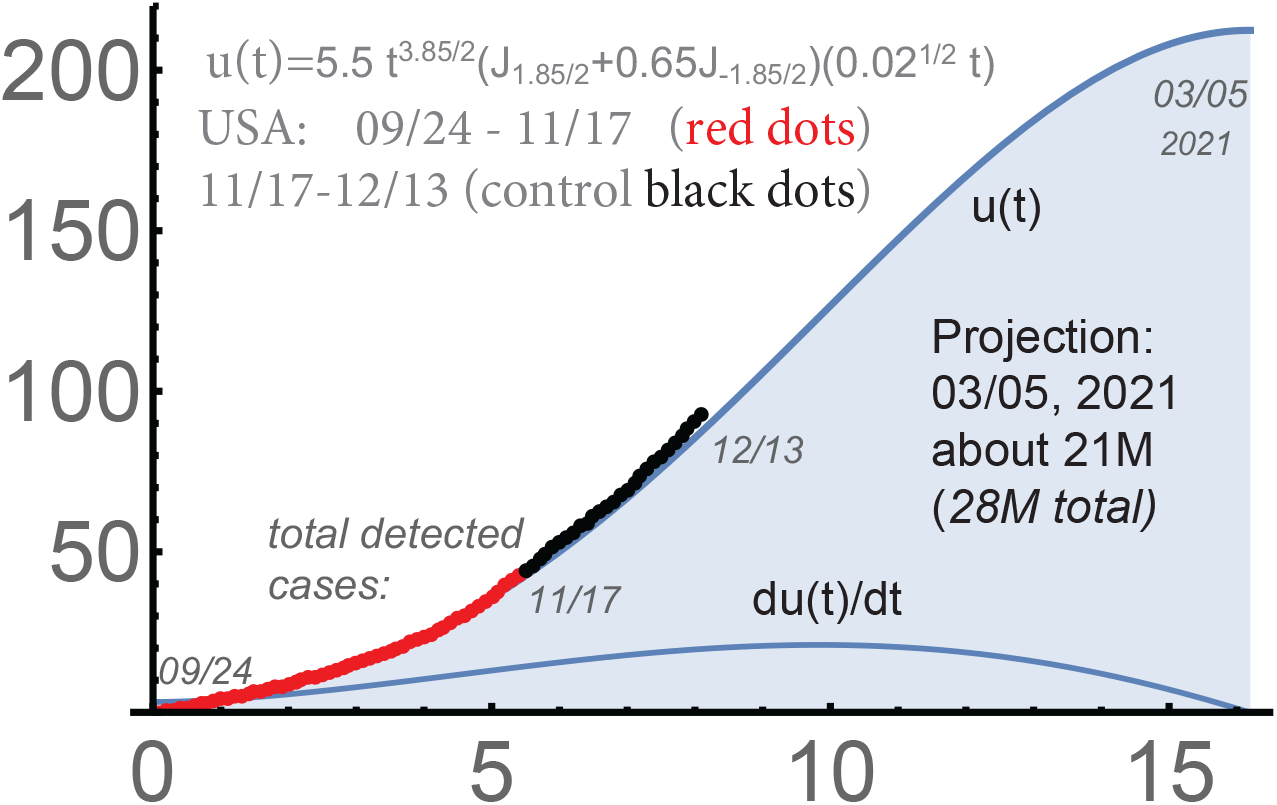
The 3rd wave in the USA.

The next control period is presented in Fig. 15. It shows that the accuracy of this forecast appeared even better in March. This is remarkable, because the previous waves in the USA were with some “unusually long” linear-type periods in the middle. Forecasting was more difficult with the USA than in almost any other countries we considered. Note that the massive vaccination began in the USA somewhat later, after the saturation on 03/05/2021: mostly from the end of March.

**Figure 15.**
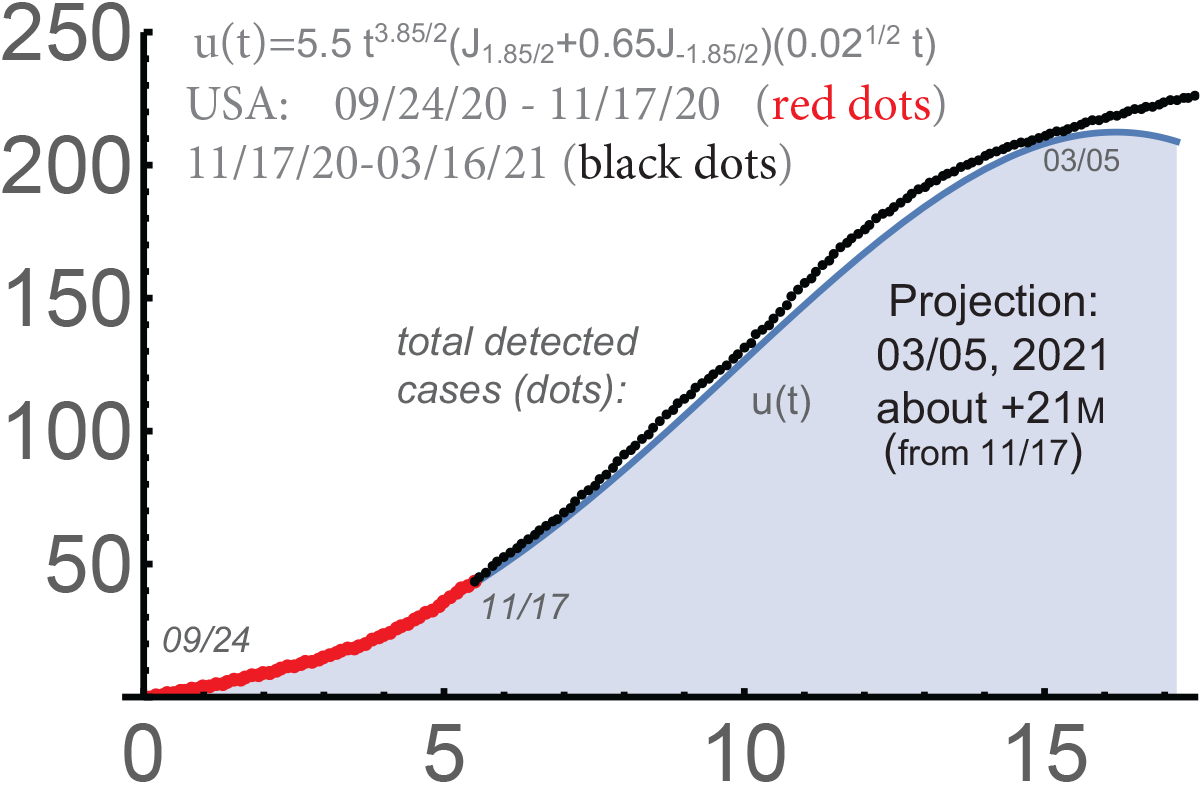
The end of the 3rd wave in the USA.

The 3rd wave in the USA was followed by some break until the 4th wave began. The 4th one was exceptionally short (for the USA). The country already reached the first stages of the herd immunity (for the current strains) in the middle of March, and the vaccination program contributed very much, as it happened in Israel, UK, etc. The current cycle of this pandemic in the USA and in quite a few other countries is hopefully on its way to the end, though this is of course only for the strains of Covid-19 that dominate now and with various reservations. The waves were “back-to-back” so far with Covid-19, so the cycles can be without significant breaks between them too.

### Delta-waves in South Africa and UK

Let us discuss the latest waves in South Africa (*ZAF*) and UK. The curves of the total number of detected cases for South Africa and UK, are surprisingly similar. There were 3 clear segments. The 1st was with relatively slow growth of the daily number of new infections, presumably because this was the period when the Delta-strain was establishing its dominance. Then it was a period of fast growth of the number of cases, actually in almost all countries with Delta-waves. It was of clear power type; the function 10(*t/*2.8)^2.9^ approximated well the number of total cases in UK for this segment. It did not last too long, and was followed by a relatively fast decline of the number of new daily cases.

In UK, the program of intensive vaccination of population in June-July and summer vacations in July-August certainly contributed to this relatively sharp decline of the number of daily cases (in spite of the relaxation of the protective measures). The reasons for this are less clear in South Africa. It is quite possible that the biology of the virus was an important factor here. Generally, viruses can be expected to lose their strength after sufficiently long sequences of transmissions; the destructive mutations are very frequent, though the proofreading feature of Covid-19 is a consideration.

It seems difficult to explain without virology why the graph of the total number of (detected) cases for the 3rd wave for South Africa (mostly “under” the Delta-strain) is so much similar to that for the previous wave there, the 2nd, and to that in UK. The shapes of the curves for UK for the 4th wave and the 3rd one are similar too. Obviously these 2 countries are in very different situations (the health care systems, the vaccinations, and other factors). So the reasons for the similarity of the parameters of the waves there can be of fundamental nature.

The similarity of the waves of infections in many so different countries remains a challenge.

There is one important reservation here: the later stages of the curves for the Delta-waves in both countries are of *significant* linear growth: the number of new daily cases became essentially constant some time after the peak, but not small. Generally, this can be an environment for new strains of Covid-19 to begin their way to dominance. These features, a sharp growth and then a sharp decline after the peak, followed by a significant number of new daily infections, can be seen in other countries experienced the Delta-waves.

In South Africa, *one u*-curve was essentially sufficient to approximate all 3 segments; Fig. 16. In UK, the spike during the 2nd segment is somewhat beyond the *u*-curve providing the optimal approximation, which is presented in Fig. 18. The projections focused on the 2nd, the most intensive, segment (until 7/15) were naturally higher; compare Fig. 18 with 19, and Fig. 16 with 17.

**Figure 16.**
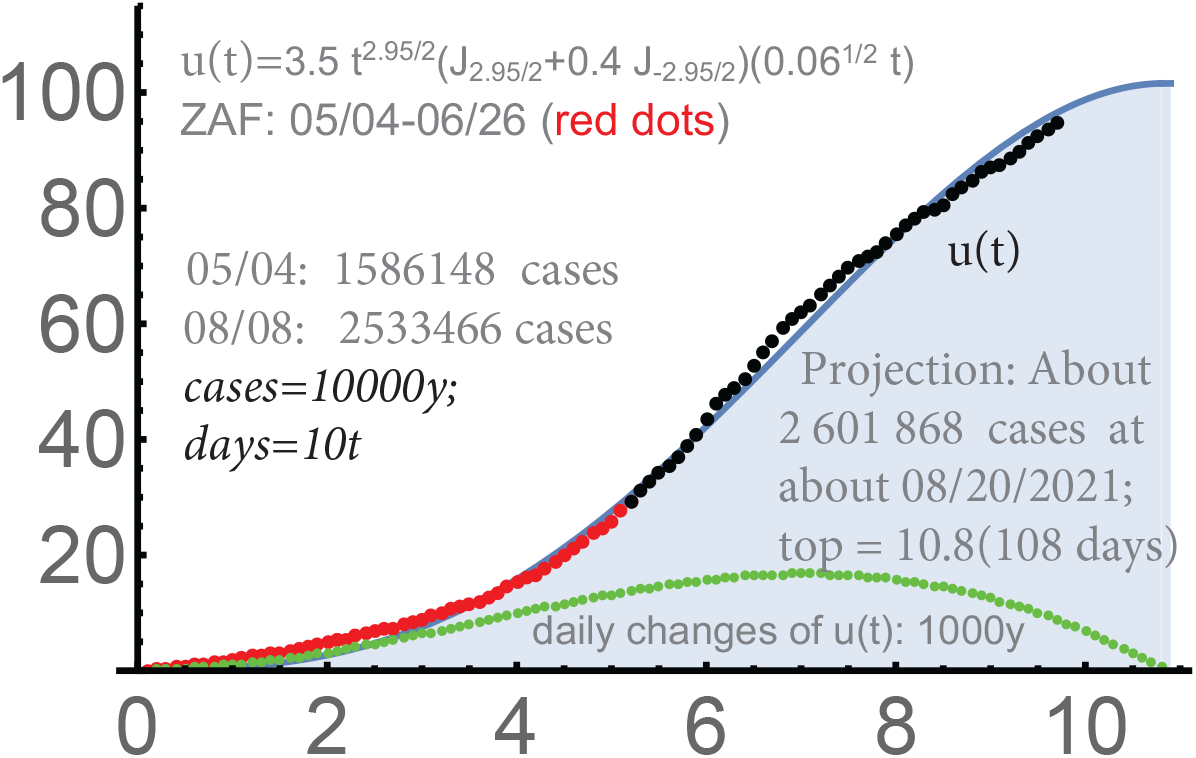
South Africa: 05/04-08/08, with the optimation for 05/04-7/15

Fig. 16 and 17 provide the projections for *ZAF* with the red dots until 06/26/2021 and until 07/15/2021. For UK, both periods were until 07/15/2021. Recall that the dots, red (when the parameters were determined) and black (the control periods), present the corresponding total numbers of detected cases diminished by that at day 1. The projections for UK and *ZAF* are provided in terms of the absolute total numbers of detected cases (from the very beginning of the epidemic).

**Figure 17.**
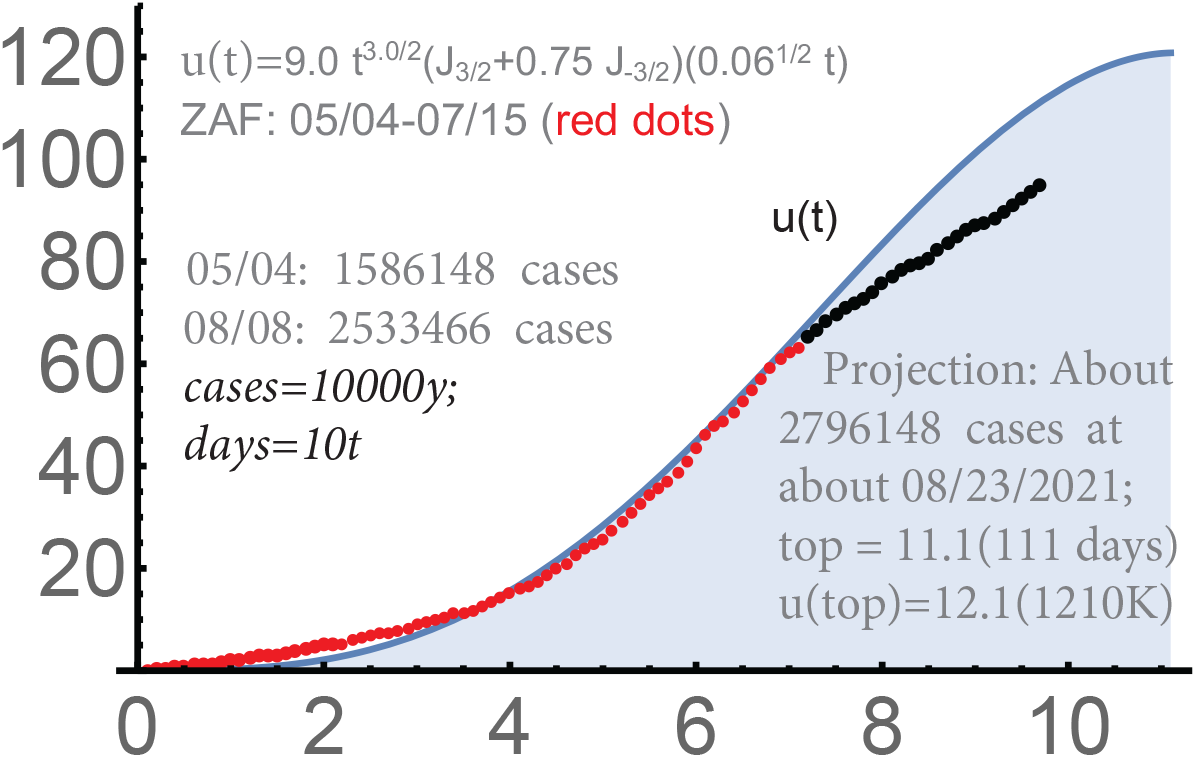
South Africa: 05/04-08/08, focus mostly on the stage from 06/15.

**Figure 18.**
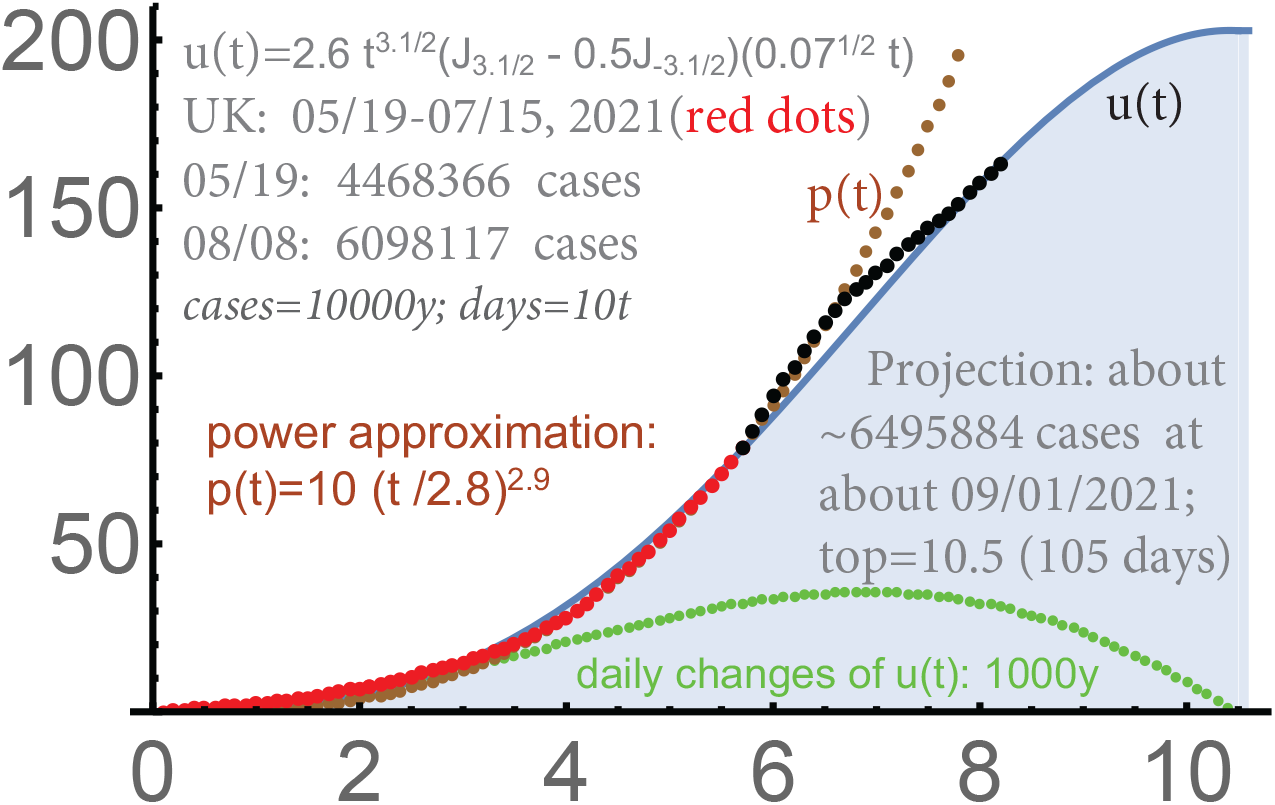
UK: 05/19-08/08, optimization for 05/19-07/15.

**Figure 19.**
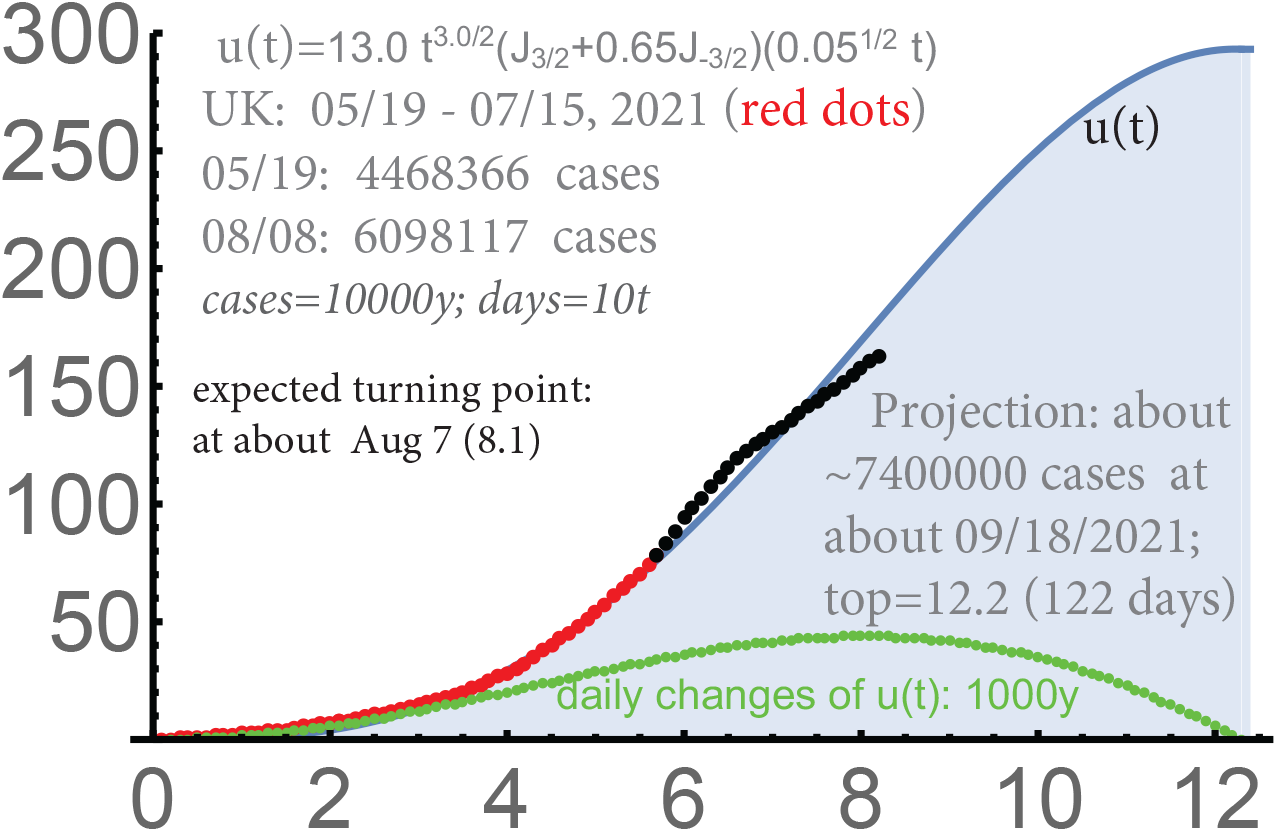
UK: 05/19-08/08, with the focus on 06/15-07/15.

### Auto-forecasting

We mostly did this for the USA and Western Europe, but any countries can be “processed” during their second phases of any waves, which under mode (*B*). Currently, there is no software for the 1st phases, i.e. the periods “under” the Bessel-type modeling.

#### The USA

`from 03 to 05, 2020`. We will provide the automated forecast for 50 states based on the period 03/17-05/27; the data were from https://github.com/nytimes/covid-19-data. Every state was processed individually `with the interaction`; see [Ch1]. Our approach to incorporating the interaction is of independent interest: we allow the curves for individual states to become `decreasing` as far as the total sum increases, which is motivated by physics.

Our program focuses on the last 20 days; however, the match with the total number of detected infections appeared `perfect` almost from 03/17/2020. See Figure 20 and compare it with Fig. 22.

**Figure 20.**
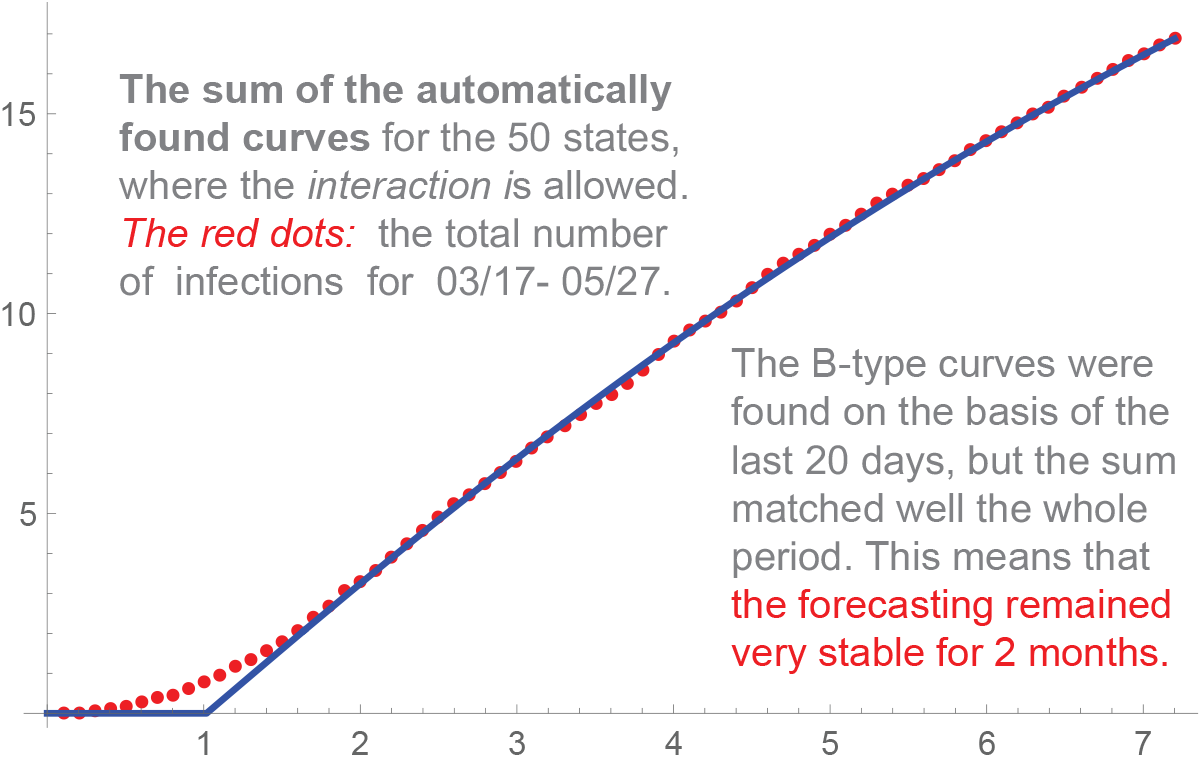
USA, the sum of the curves for individual states.

Such a high level of stability is rare in any forecasting, which made the chances good for reaching the saturation around 9/19/2020. This was our projection based on Fig. 20 and on the assumption that the level of protective measures would remain essentially unchanged. Recall that the saturation for phase 1 is of technical nature: it does not mean the end of the wave. Normally, it is followed by a period of modest linear growth of the total number of infections, which we model using *u*_*B*_(*t*); this is the 2nd phase. Also, there are always remaining and new clusters of infection and no country is really isolated. In some countries we analyzed, mode (*A*) alone was almost sufficient to model a wave until its end in several countries. However mode (*B*) (and the second phases) were mostly present,

Concerning the projection 9/19/2020 in the USA, the hard measures were significantly reduced there at the end of May practically in all 50 states. As a result, the number of states that reached phase 2 dropped from about 22 at 5/27 to 8 at 7/12 (2020). Then, in the second half of June, the USA entered the `second wave`.

Similarly, the program was quite stable for phase 2 for the 2nd wave in the USA … before it entered the 3rd wave in the middle of September of 2020. We recall that the program is written for the 2nd phase only, when constant (daily) monitoring and tuning the process of management is more relevant. This is the period when the hard measures are being reduced and the constant adjustment are needed. However *a posteriori*, using the data for the whole wave, we never had difficulties with finding the parameters *b, d* for the *whole* second phase.

#### Europe

`summer 2020`. The situation was quite stable in Europe in summer of 2020. We provide in Figure 21 a sample forecast our automated system produced for Western Europe `till the end of July, 2020`. It was for 45 countries.

**Figure 21.**
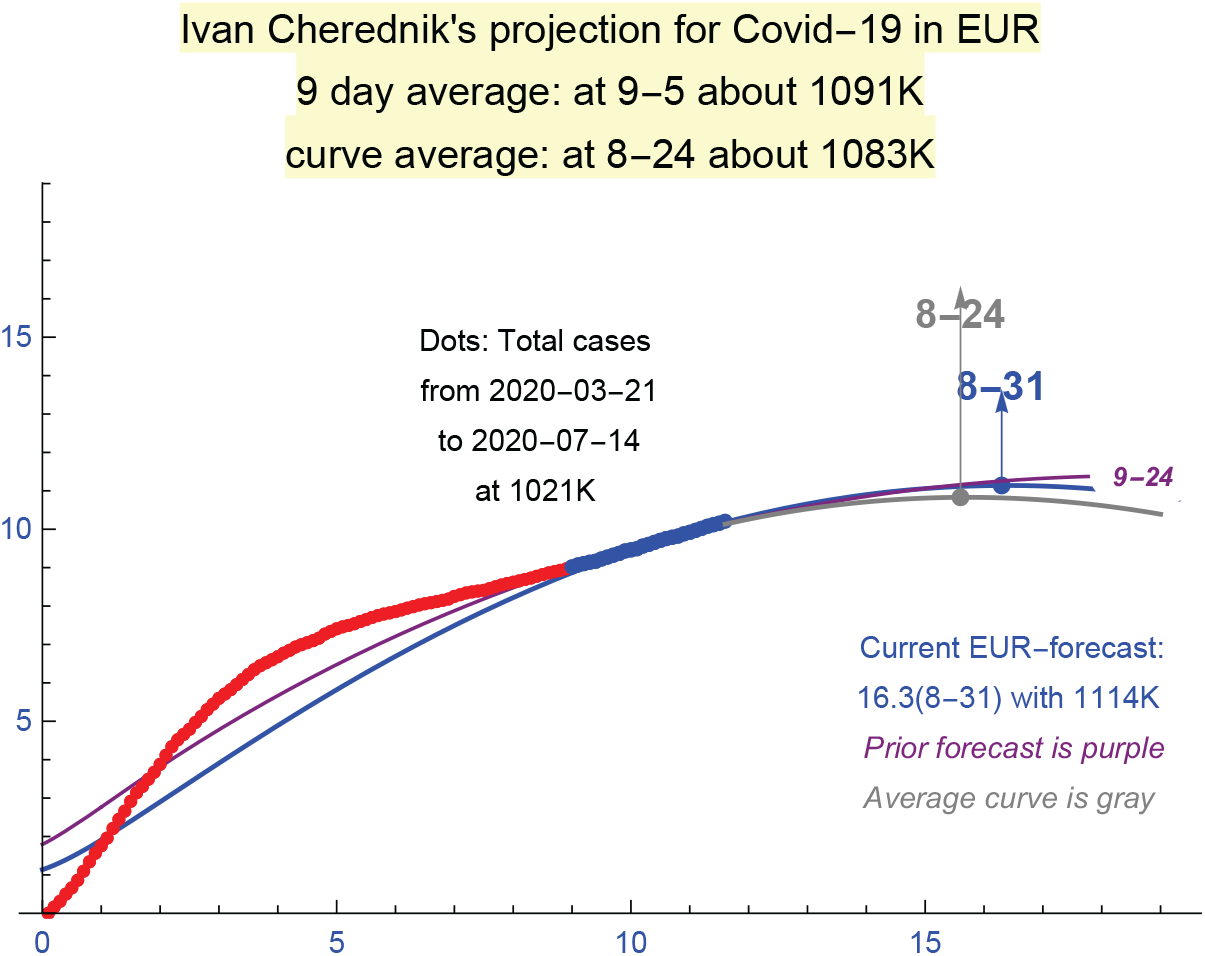
An auto-forecast for Europe as of 7/14 (2020)

Here and below the *curve average* is the maximum and the corresponding value of the average of the 9 last curves *u*_*B*_(*t*) for the country or the region. I.e. we consider the average of 9 curves we obtain for 9 consecutive days and then determine its maximum. The *9-day average* is the simple average of the corresponding maxima for these 9 curves; i.e. it is the moving average. The main source of Covid-19 data we used was: https://ourworldindata.org/coronavirus.

As of July 8, 2020, the forecasts were sufficiently stable, though Sweden, Poland, Portugal and some other countries did not reach phase 2 at that time. Such stability changed in fall 2020 due to the end of the vacation periods and the beginning of the school year.

`Summer 2021`. Fig. 22 provides an auto-generated projection for the USA as of 06/09/2021. This is without the state-by-state analysis: the total numbers of detected infections for the whole country are used. The projections are supposed to be constantly renewed during phase 2. The fact that the curve of (actual) detected cases was very well approximated by our *u*_*B*_(*t*) for the whole period of 90 days is very remarkable. Only the last 20 days are used for finding the (current) parameters.

**Figure 22.**
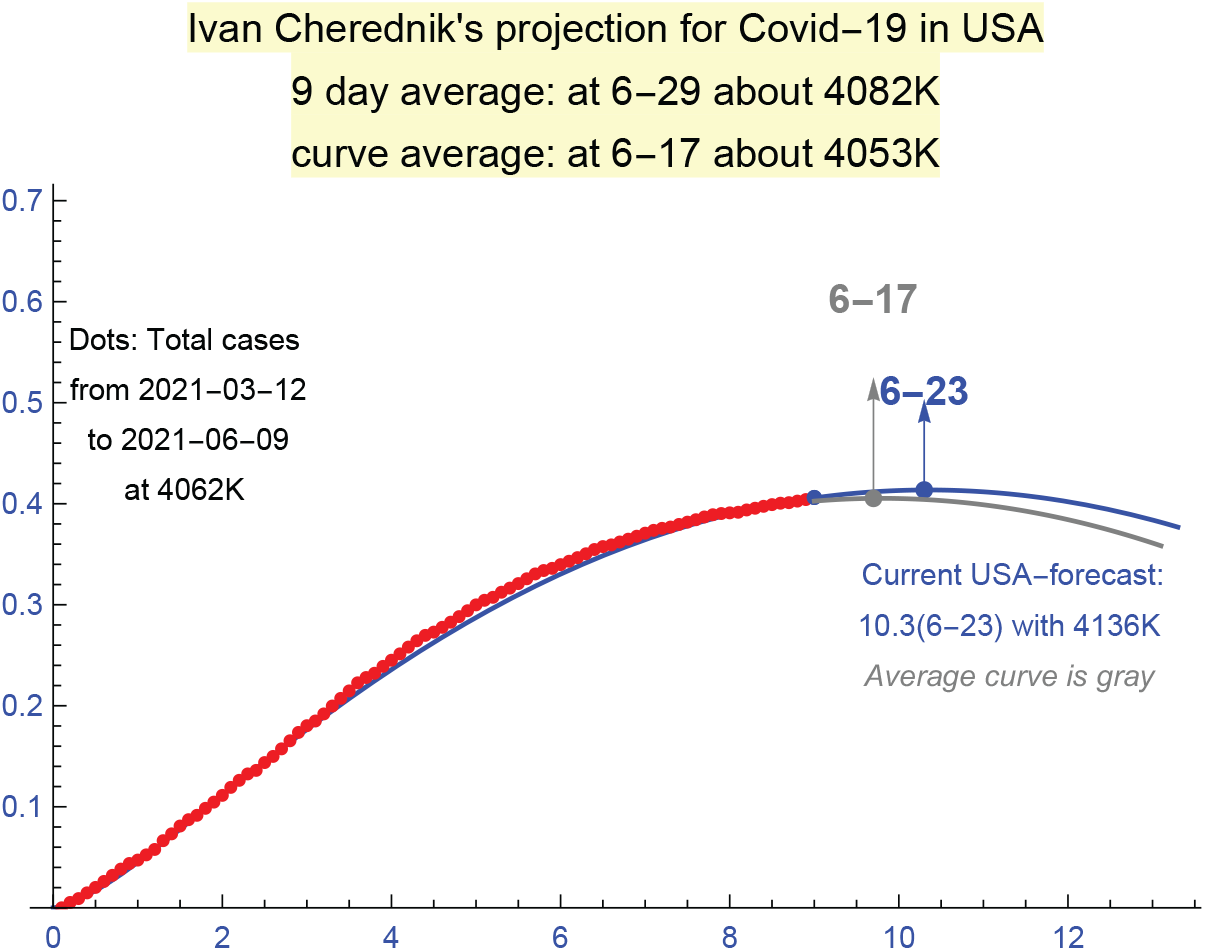
A sample forecast in the USA: 06/09/2021.

This is a strong confirmation of our `two-phase solution`. The USA was in the middle of the intensive vaccination program during this period. Also, it was already close to the initial levels of herd immunity (for the current strains) in March-May. The aggressive vaccination program and continued restrictions of contacts (though significantly relaxed in May-June) influenced this period.

For Europe during summer 2021, a similar projection is provided in Fig. 23. As with the USA, a perfect match with the actual data is for the whole period of 90 days; only the last 20 days are used to find the parameters so this is an indication of the stability of such forecasts.

**Figure 23.**
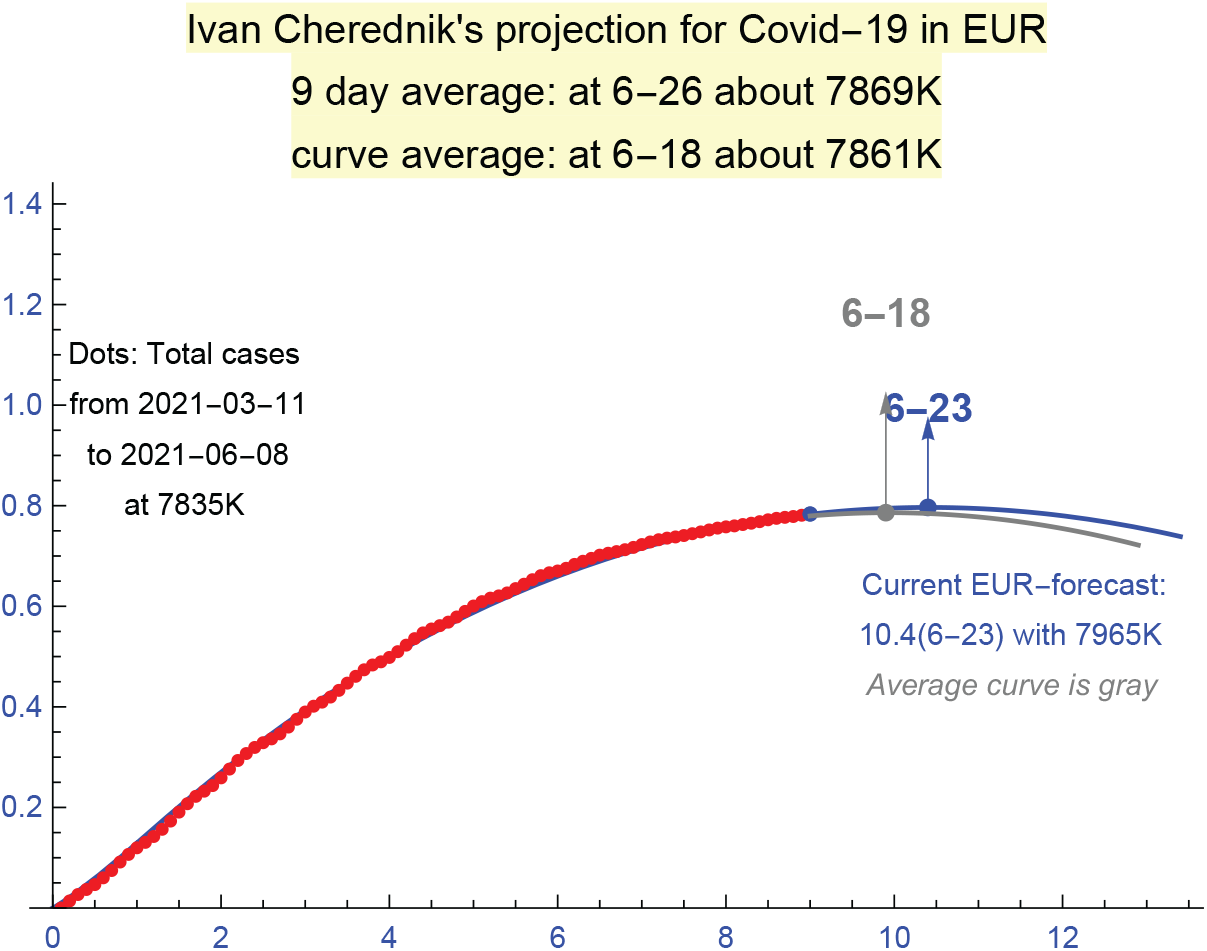
A sample forecast in Europe: 06/08/2021.

The growth of the number of the cases is expected in Fall 2021 due to the impact of the Delta-strain and the end of summer vacations. This strain already heavily influenced UK, South Africa and a growing list of other countries.

The Delta-strain can be some transition between cycles. However more significant modifications and recombinations of the existing strains are more likely to dominate the next cycle. Really new strains can be expected to emerge, including those “responding” to the vaccination programs.

The situation with Covid-19 remains far from “normal” in many countries. However the levels of herd immunity for the current strains reach sufficiently high levels and there are intensive vaccination programs in quite a few countries. These factors can be expected to end the current cycle, but this is of course conditional. Much will depend on the evolution of the strains of Covid-19 and there are already some new waves with a potential to trigger a new global cycle, which can be close-by. The “classical” seasonal periodicity of waves/cycles is not really applicable to Covid-19.

Obviously, a combination of protective measures, herd immunity, and the vaccination programs began to work. According to the latest data, the hard measures remain the key factor in controlling the spread of Covid-19; the periodic vaccination of the population is such, but this alone seems insufficient without other protective measures, including the self-imposed ones. We did not really observe exponential growth of the total number of cases (unless during short initial periods), but unrestricted power growth with exponents *c >* 2 is very devastating. Detection-isolation-tracing, the classical complex of hard measures, remains of great importance even upon the extensive vaccination.

Hopefully we will be better prepared to new cycles of Covid-19, including improved mathematical tool for forecasting the spread of epidemics, better understanding the uniformity of the waves of the infections, automated methods for finding the parameters that determine the spread. The purpose is of course to making solid mathematical predictions for the durations and magnitudes of the waves of infections.

### Summary

Modeling Covid-19 appeared quite a challenge for existing mathematical methods, which are mostly based on the `SID`-type approach, suggested in the early 20th century. The following features of Covid-19 obviously require new methods.

1. The curves of total numbers of detected infections are mostly of power-type for Covid-19, where the exponent diminishes over time.
2. The saturation of the initial waves of Covid-19 is mostly because of the protective measures, not due to the herd immunity.
3. The range and intensity of the protective measures used to fight Covid-19 are exceptional in the history of epidemics.

These factors are not really new in epidemics, but they do require a new mathematical theory. The prior approaches appeared insufficient for modeling the spread of Covid-19. For instance, the power-type growth of the total number of infections cannot be addressed within the `SID`-type models.

Our theory seems the first one when the power growth of the spread and the active epidemic management, including self-imposed restrictions, are considered the major factors. These assumptions result in differential equations depending only on the initial transmission rate and the intensity of the protective measures. These parameters make perfect sense theoretically and practically, and can be measured reliably during relatively early stages of the waves of Covid-19.

The actual graphs of the total numbers of detected cases in many countries (all we considered) are described uniformly and with surprisingly high accuracy by our curves. The `2-phase solution` is a combination of the Bessel-type curve for phase 1, which is the key in our approach, and its certain version for phase 2 (in terms of elementary functions). We note that the corresponding saturations were mostly due to the active management till summer 2021. They are of unstable nature: reducing the protective measures may result in the recurrence. Modeling and forecasting this kind of saturations requires sharp mathematical tools, which we hopefully provide.

## Methods

The starting point of our approach to modeling the total number of infections during epidemics is the `power growth hypothesis`, which has solid confirmations with Covid-19, practically in all countries (apart of small initial periods). This is without considering epidemic management; we deduce it from the principle of `local herd immunity`.

The saturation of the corresponding waves of the spread of Covid-19 within this cycle (at least) is mainly due to the `protective measures`, the process of active management of the epidemic including our own actions. Protective measures are of course not unique for Covid-19, but their range and intensity reached unprecedented levels. Our model connects this kind of saturation with the asymptotic `periodicity of Bessel functions`, one of the deepest results in their theory.

This is very different from the classical approaches based of SID, SIR, SIER models and their variants, and the approaches in the neighboring segments of ecology: `invasion and interaction between species`. The saturation for us is a dynamic equilibrium between the virus invasion and our protective measures, including very important self-imposed ones. This is not because of the classical herd immunity and is actually parallel to invasion ecology.

Due to a very limited number of parameters, actually 3 for our `two-phase solution`, our model is much more rigid than any other ones. The curves we obtain match very well the actual graphs of the total numbers of infections practically in all countries we examined (many). These parameters are quite meaningful mathematically and epidemiologically. The key are *c*, `the initial transmission rate`, and *a*, `the intensity of hard protective measures`, which can be determined reliably at relatively early stages of particular waves of the epidemic.

Concerning forecasting the waves, it can be reduced to “predicting” *c, a, C* and the coefficient of the non-dominant solution *u*^2^ of (1). The challenge is to do this at early stages of the waves. We can do this “manually”, but deep machine learning is natural here.

Our parameters are generally different for different waves, but we see some patterns here. The second waves are almost always for somewhat higher *c* and significantly lower *a*, the intensity of hard protective measures. The 2nd wave in India was exceptional: *c* dropped and *a* increased. The number of contacts was reduced faster during the 2nd wave, much broader than the 1st one, and the protective measures, including important self-imposed restrictions, were significantly stronger. As for the 1st wave, the match with our *u*(*t*) was almost perfect.

We obtain a very good match `practically for the whole periods` of the waves of Covid-19 in many countries. It is actually surprising for such stochastic processes as epidemics, and of course this is a strong confirmation of our assumptions. In spite of various simplifications, it appeared that equations (1) and (2) capture very well the dynamic of the waves of Covid-19. We present our motivation for their usage for modeling epidemics, however the key for this surprising discovery can be in the universality of these equations.

Since our theory was created and posted in the middle of April 2020, which was mostly in the middle of the 1st waves of Covid-19, we had a unique opportunity to test it in the course of epidemics. Namely, we systematically determined the parameters of our curves during relatively early stages and then tested the corresponding projections extensively for sufficiently long `control periods`.

Our usage of control periods is similar to routine testing the quality of the models used for forecasting share-prices in stock markets, where no approach can be accepted without real-time runs and carefully crafted historic experiments that exclude any “usage of future” as far as possible. This kind of “discipline” is not present in forecasting the epidemics, at least by now.

The results of checking our theory during the control periods, including the outputs of the automated forecasting programs we developed, are an important part of papers [Ch1, Ch3].

An example of a long term forecast is the 3rd wave in the USA discussed above. It shows that potentially our tools can be used to obtain reasonable projections at relatively early stages of the waves.

## Conclusion

We demonstrated that Bessel-type functions describe very well the growth of the total number of detected cases in many countries. Mathematically, we successfully model the passage from ∼ *t*^*c*^, describing the initial growth of the total number of detected cases, to ∼ *t* near the turning point, when the number of new (daily) cases stabilizes, and then almost all the way to the saturation of the current wave. This is mode (*A*).

At the late stages of the waves of Covid-19, we generally switch to mode (*B*), the differential equations describing more relaxed management (soft measures begin to dominates); their solutions are in terms of elementary functions.

Here *c* is the `initial transmission rate`, which can be captured at relatively early stages of the waves of Covid-19, though it is more reliable to determine it near the turning point. It remains the same for both modes, (*A*) and (*B*).

Our differential equations describe the epidemics under active management, the system of protective measures, where hard measures play the key role. The self-imposed restrictions on the contacts during the epidemic are and always were important protective measures. Constant receiving reliable information on the course of the epidemic by the population is of obvious significance here. The travel restrictions, closing the places where the spread is the most likely, and the vaccination are classical state controlled measures.

The saturation due to active protection measures is of unstable nature, unless the herd immunity (for the current strains) becomes a significant factor. Its forecasting requires an exact mathematical theory, which we try to provide. There will be an endless discussion of the efficiency of different measures and different management approaches until verifiable trustworthy mathematical models and the corresponding software are developed and implemented practically.

The verification of any models, including this one, does require algorithms that can be used by anyone, not only by their creators, the ultimate test of their validity. This is one of the reasons why we wrote our programs. They are posted in [Ch1, Ch3] and can be used by anyone to model any countries and regions, though only for the late stages of Covid-19 so far (mode (*B*)).

The new theory seems a solid basis for reaching the next level, which is forecasting. It already describes the curves of total numbers of detected infections with high accuracy and with surprisingly high level of forecasting and stability of the auto-projections for later phases. Though forecasting is always a challenge.

### Basic parameters

The small number of the parameter we employ explains well the uniformity of the curves of total numbers of detected infections of Covid-19 in many so different countries, as well as the mathematical similarity of different waves. They are:

1. the initial transmission rate *c*, which can be determined at relatively early stages of the current wave,
2. the intensity of hard measures *a*, which become sufficiently stable near and after the turning point,
3. the intensity *b* of the measures during the second phase, which begins near the end of phase 1.

In contract to *c*, the intensity of the measures is of course more time-dependent, but *a* appeared sufficiently stable for long periods. Concerning *b*, it must be adjusted constantly at the later stages, because no country is really isolated and the management of the epidemic, including the self-imposed protection measures, becomes more flexible at these stages, so less predictable mathematically.

The scaling coefficient *C* of *u*(*t*) is adjusted to match the real numbers of cases. The coefficient of *u*^2^, the non-dominant solution of our system of ODE, is another parameter. It is mainly responsible for the “effects of the second order” and does not seem really critical for forecasting; the dominant Bessel-type solution *u*^1^ is expected to be sufficient for this. There are technical reasons why *C* increases when *u*^2^ is added with a significant positive coefficient: *u*^2^(*t*) becomes negative for sufficiently large *t* and contributes greatly for such *t*.

The coefficient *C* is generally a technicality, to make the output convenient to present as a graph. However it provides valuable information when different waves of the epidemic are compared in the same country. An example is our analysis of the 1st and 2nd waves in India. The whole country was affected during the 2nd wave, when only some strata of this huge and diverse country were mostly infected during the 1st wave. The coefficient *C* dramatically increased during the 2nd wave there, when the coefficients *c, a* became similar to those in Europe during the 1st-2nd waves. The match of our *u*(*t*) and the total number of detected cases was very good with the 2nd waves.

Mathematically, *u*^2^ to captures some features of the curves of total numbers of detected infections, especially during the initial stages. It can be “seen” in quite a few countries, though *u*^2^ mostly influences the effects of second order. From the view point of our approach, *u*^1^ and *u*^2^, both, could be expected to occur, which was really confirmed. See [Ch1] for some discussion.

The fact that we were able to describe such complex stochastic processes as epidemics with only 3 basic parameters seems a real discovery. This worked very well almost everywhere (in all countries we considered), but it will take time to understand the scope of this new theory and to begin using it practically. Obviously such a remarkable universality can have deep roots in general biology and the theory of random processes. The same system of ODE serves quite a different situations, including momentum trading in stock markets. It models `MRT`, momentum risk-taking. Its “deduction” is different for epidemics (and in invasion ecology). The price function in momentum trading is replaced by the `protection function` *p*(*t*) for (1).

### Beyond this paper

Needless to say that the vaccinations makes us closer to the herd immunity for the current strains of Covid-19. There are many challenges here [AVTC], and, anyway, the control of the efficiency of the vaccination programs requires exact mathematical tools; there are other factors. In our approach, this means measuring their impact on the parameters, especially on *c*, the transmission rate.

We hopefully approach the end of the 1st cycle in the Northern Hemisphere within the current dominant strains, though this is far from certain. Basically, the end of a `cycle` is when the current strains become less infectious due to growing immunity, vaccinations, the change of the season and similar factors. The epidemic can continue between the cycles in other places, zones, and zoonotic reservoirs, and a significant renewal of the strains can be expected in new cycles. Covid-19 demonstrated a unique ability to produce many ways one after another, sometimes in the very same areas; further developments of the current strains can be expected and their recombinations.

Concerning the new strains, presumably about 6 months are necessary for them to reach a significant presence counting from their “first appearance”. The latter can be the moment of their creation, which can be expected at late stages of the prior waves, or when new strains are “imported” from other regions. Also, the immunity upon the recovery from Covid-19 or upon the vaccination is expected to begin to fade during approximately 6 months, which is an important factor.

Covid-19 demonstrated a robust potential for creating various `VOC`, variants of concern. Its ability to evolve resisting the vaccines and the treatment remains to be seen, but it can be expected to be ample. Generally, it can take only several infected individuals where the virus can “stay” for 1-3 months to generated many mutations, but fortunately very few of them are stable and have a chance to reach any dominance. The protective measures greatly restrict their spread (always did); a mathematical theory of their impact combined with `power-law of epidemics` is the purpose of [Ch1, Ch3] and this paper.

The presence of weakened viruses due to sufficiently long sequences of transmissions works in the same direction. This mechanism is likely to clarify: (*a*) great uniformity of the curves of total cases, and (*b*) their saturation well before any herd immunity is reached. However more biological research is needed: how the mutations weaken the virus and how transmissible the weakened viruses can be. The proofreading feature of Covid-19 is of importance here, but weakening the virus due to mutations of destructive type can be one of the key factors.

A natural challenge is to understand the formation of waves within one cycle. Recall that we stop the usage of Bessel functions when the corresponding *u*(*t*) begins to diminish, which is impossible for the `total` number of cases our *u*-functions describe. Then we switch to mode (*B*) and use *u*_*B*_(*t*) till its own maximum.

Given a country, the waves and their periodicity certainly follow some patterns. However the pauses between them and the parameters *c, a, C* fluctuate significantly. If the next wave is due to the relaxation of the protective measures at the end of the previous one then the problem can be addressed mathematically: the changes of the main parameters have some tendencies. However there can be many other factors influencing new waves, including new strains. Anyway, 2-4 waves (in one country) are insufficient for modeling their recurrence and the corresponding evolution of the parameters.

### Main practical applications

They seem for modeling individual waves. Namely, the natural aims are as follow:

1. Forecasting the duration and the amplitude of the waves of Covid-19 and similar epidemics of “power-type”; the main parameters can be reliably determined near or even before the turning points of the spread.
2. Creating software for monitoring the dynamic of the epidemic. This is especially needed at the later stages of its waves (2nd phases in our terminology), when tuning the management becomes important.
3. When comparing different waves in the same country, conclusions can be made about the changes in the epidemic management (including self-imposed restrictions), and the changes of the virus behavior.
4. Extending this theory from modeling epidemics to Invasion Ecology, more specifically, to transient processes of the invasion and interaction between 2-3 species; applying it to momentum risk-taking.

## Supporting information

forecasting program for later stages

## Data Availability

“A surprising formula for the spread of Covid-19 under aggressive management”, MedRxiv, doi: 10.1101/2020.04.29.2008448 , 2020.

https://www.medrxiv.org/content/10.1101/2020.11.20.20235903v2

## Acknowledgements

The author thanks very much David Kazhdan for valuable comments and suggestions; a good portion of this paper presents author’s attempts to answer his questions. Many thanks to Eric Opdam and Alexei Borodin for their kind interest and ETH-ITS for outstanding hospitality; special thanks are to Giovanni Felder and Rahul Pandharipande. We acknowledge support by NSF grant DMS– 1901796 and the Simons Foundation.

