## Supplementary material for "Modeling the waves of Covid-19": forecasting program for later stages: README-WORLD.pdf

USE THIS PROGRAM, CREATED BY IVAN CHEREDNIK, AT YOUR OWN RISK. ONE PROJECTIONS IS ON THE BASIS OF THE LATEST 20 DAYS. THE AVRG CURVE IS THE AVERAGE OF 9 PRIOR FORECAST CURVES. EXPECT THE "SATURATION" TO MOVE OVER TIME. IF THERE IS NO SATURATION THEN THE FORECAST IS FOR THE 4 NEXT MONTHS. THIS PROGRAM IS DESIGNED FOR THE LATER STAGES OF THE WAVES.

TO USE IT, YOU MUST AGREE WITH THESE TERMS. ALL CREDITS GO TO IVAN CHEREDNIK, THE DATA: githubusercontent.com/owid/covid-19-data. THE PROGRAM WAS CHECKED FOR MATHEMATICA 11.

1) PUT THE FILES IN ANY FOLDER. IF YOU USE MathKern, THEN MAKE IT THE START FOLDER IN "MathKern Properties". ALTERNATIVELY, AND FOR "FULL" MATHEMATICA, BEGIN WITH COMMAND SetDirectory["your folder"], NOTICE QUOTES. THEN DO "<<forwor.txt"; THE OUTPUT WILL BE IN "world-full.pdf" (IF YOU USE MathKern).

2) EXE-TOOL. CHANGE PATH OF "math.exe" TO ITS ACTUAL ONE IN YOUR SYSTEM, WHICH IS IN "mathpath.txt" (WITH QUOTES THERE). CLICK ON "foreurun.exe", AND AFTER 2-3 MINS THE FILE "world-now.pdf" WILL OPEN. COVID DATA ARE UPDATED DAILY; THERE IS NO HARM TO RUN "foreurun.exe" AS MANY TIMES AS YOU WISH.

3) FILE "worldfile.txt" CONTROLS COUNTRIES AND REGIONS. MAKE reg="ALL" FOR THE WORLD, IT CAN BE reg="Europe","North America",ETC. BY MAKING grp=1 YOU WILL RUN IT FOR YOUR GROUP grpx={"United States","Brazil"}, ETC. MAKE delprev=1 FOR THE 1st RUN WITH A NEW GROUP/REGION: OLD OUTPUTS WILL BE DELETED.

THE PDF FILES PRODUCED BY "forwor.txt" AND ITS OUTPUT IN MATHEMATICA CONTAIN FURTHER RELEVANT INFORMATION. THIS SOFTWARE IS NOT FOR ANY COMMERCIAL USE; THIS IS A RESEARCH TOOL! YOUR COMMENTS ARE APPRECIATED. -IVAN
